# The engineered monoclonal antibody tobevibart enhances HBsAg capture by Fc receptor positive cells leading to activation of HBV-specific T cells

**DOI:** 10.1101/2025.01.13.25320453

**Authors:** Lucia Vincenzetti, Rachel Wong, Roberta Marzi, Barbara Guarino, Erin Stefanutti, Sneha V. Gupta, Laura E. Rosen, David M. Belnap, Li Wang, Yi-Pei Chen, Julia di Iulio, Amin Momin, Karen E. Tracy, Sharvari Deshpande, John M. Errico, Nicole Sprugasci, Alessia Peter, Lillian Seu, Daniel Cloutier, Chin H. Tay, Gyorgy Snell, Nadine Czudnochowski, Florian A. Lempp, Colin Havenar-Daughton, Fabio Benigni, Antonio Lanzavecchia, Kosh Agarwal, Man-Fung Yuen, Heiner Wedemeyer, Ed Gane, Ann Arvin, Davide Corti, Michael A. Schmid

**Author notes:** **Correspondence:** Michael A. Schmid via. **Clinical trial:** VIR-3434-1002, phase 1, ClinicalTrials.gov identifier NCT04423393.

## Abstract

**Background & Aims:** Immune targeting is likely required for functional cure of chronic hepatitis B (CHB). Tobevibart, a human monoclonal antibody against hepatitis B virus (HBV) surface antigen (HBsAg), neutralizes HBV and hepatitis delta virus (HDV). This study aimed to characterize effects of the engineered GAALIE Fc of tobevibart on HBV immune responses.

**Methods:** We studied tobevibart and its equivalent HBC34*-GAALIE *in vitro* using electron microscopy, FcγR reporter cells, and primary human or mouse immune cells to assess HBsAg binding, dendritic cell (DC) activation, and T cell stimulation. Tobevibart-mediated binding of HBsAg to immune cells was evaluated also in a phase 1 clinical trial in patients with CHB.

**Results:** The GAALIE Fc of tobevibart mediated gain of function in FcγR signaling in immune complexes (ICs) with HBsAg compared to wild-type (WT) Fc and increased binding of HBsAg to neutrophils and monocytes *in vitro*. Similarly, dosing of 300 mg tobevibart in patients with CHB mediated binding of HBsAg to these cells *in vivo*, concomitant with reducing HBsAg in circulation. *In vitro*, ICs of HBC34*-GAALIE and HBsAg activated human DCs significantly more than HBC34*-WT. These DCs presented antigen and stimulated HBsAg-specific human T cells. Similarly, ICs of HBC34*-GAALIE and HBsAg activated DCs from mice transgenic for human FcγRs and stimulated CD4+ T cells from immunized animals more than ICs with HBC34*-WT.

**Conclusions:** We demonstrate that tobevibart combines the advantages of potent neutralization of HBV and HDV, FcγR-mediated reduction of HBsAg, and Fc-dependent enhancement of T cell responses. Tobevibart is currently under clinical investigation alone or in combination with other agents to treat patients with chronic hepatitis delta and to induce functional cure of patients with CHB.

**Impact and Implications:** Chronic infection with hepatitis B virus (HBV) or co-infection with hepatitis delta virus (HDV) can cause severe liver disease and cancer. We previously showed that the monoclonal antibody tobevibart potently neutralizes HBV and HDV. Here we show that the engineered Fc region of tobevibart effectively interacted with several immune cell types *in vitro*, which may support the rapid removal of damaging virus proteins from circulation, potentially activating T cell responses that may control HBV infection long term.

## Introduction

Hepatitis B virus (HBV) infection can cause self-limiting acute hepatitis or lead to chronic hepatitis B (CHB), potentially progressing to liver fibrosis, cirrhosis and hepatocellular carcinoma. Worldwide, CHB causes 1.1 million deaths annually [1]. Co-infection with hepatitis delta virus (HDV), which uses HBV surface antigen (HBsAg) for its own envelopment, causes an even more severe form of viral hepatitis.

Acutely infected individuals who resolve HBV infection develop adaptive immunity, including CD8+ T cells that kill infected hepatocytes, CD4+ T cells that provide help to CD8+ T cells, and B cells that produce neutralizing anti-HBs antibodies [2]. In contrast, immune evasion mechanisms, such as limited innate recognition of HBV and prolonged exposure to high levels of HBsAg subviral particles (SVPs), can create a tolerogenic environment that results in adaptive immune exhaustion and chronic HBV replication in the liver [3].

The current goals of CHB treatment are to reverse immune exhaustion and induce functional cure, defined as persistent loss of serum HBsAg and HBV DNA after a finite course of treatment. To achieve functional cure, induction of HBV-specific T and B cell immune responses is likely essential. The currently approved HBV therapeutics, nucleos(t)ide analogues (NAs), that suppress viral replication must be administered daily and do not reduce HBsAg levels. Combining NAs with pegylated-interferon-α (PEG-IFNα) only results in low rates of HBsAg loss [4]. An alternative approach is the use of HBsAg-specific (anti-HBs) antibodies to clear HBsAg. Polyclonal hepatitis B immunoglobulin (HBIG), purified from serum of vaccinees, is effective prophylaxis for neonates born to HBV-infected mothers or liver transplant recipients. However, HBIG and several anti-HBs monoclonal antibodies (mAbs) with a wild-type (WT) Fc portion reduced HBsAg levels only transiently when administered therapeutically [5], and, thus, HBIG is not useful for treating CHB.

Achieving CHB functional cure will likely require therapies with multiple modes of action, whether as a single drug or a combination of drugs, to mediate i) virus suppression, ii) reduction of HBsAg, and iii) induction of long-acting HBV-specific T and B cell responses. We previously described the direct antiviral modes of action of the human anti-HBs mAb tobevibart (VIR-3434) that was discovered from the blood of an HBV vaccinee, was Fc engineered, and is currently in clinical development for treatment of CHB and chronic hepatitis D (CHD) [6]. Tobevibart binds to a conserved discontinuous epitope within the antigenic loop of all forms of HBsAg (small, medium, large) and neutralizes all genotypes of HBV and HDV *in vitro*. In human liver-chimeric mice, tobevibart efficiently blocked intrahepatic viral spread and reduced HBsAg as well as HBV and HDV viremia [6]. Beyond virus neutralization, indirect mechanisms mediated by the Fc domain, which interacts with Fc gamma receptors (FcγRs), have previously been shown for other anti-HBs mAbs to reduce HBsAg [7, 8] and induce adaptive T and B cell responses in mice [7, 9, 10].

To further enhance Fc functions, tobevibart was engineered to extend its half-life by the (M428L, N434S) “LS” mutation [11] and to increase effector functions by the (G236A, A330L, I332E) “GAALIE” mutation, which increases binding to activating FcγRs IIa and IIIa and decreases binding to inhibitory FcγRIIb [12]. In a melanoma model, a mAb with GAALIE Fc modification reduced lung metastasis more than a mAb with wild-type (WT) Fc in mice transgenic for human FcγRs (huFcγR mice) [12, 13]. For SARS-CoV-2 infection, two mAbs with the GAALIE Fc protected huFcγR mice against weight loss and death more effectively than the same mAbs with WT Fcs [14]. A broadly neutralizing mAb targeting the haemagglutinin stem of influenza virus and engineered with GAALIE Fc was more protective in huFcγR mice, activated classical DCs (cDC), and primed naïve CD8+ T cells more rapidly compared to the same mAb with WT Fc [15]. This antibody-mediated increase of T cell responses has been referred to as a “vaccinal effect” because antigen-specific T cells are induced or amplified by Fc-dependent enhancement of antigen presentation, similar to a vaccine response.

Here, we characterize the Fc-mediated functions of tobevibart by using electron microscopy, reporter cell assays for FcγR signaling, primary human cells, including in patients dosed with tobevibart, and a mouse model transgenic for human FcγRs. We showed that tobevibart induced short-term effector functions that can contribute to HBsAg removal from circulation and has the potential to stimulate long-lasting HBsAg-specific T cell immunity.

## Results

### Tobevibart cross-linked HBsAg subviral particles (SVPs) and formed immune complexes (ICs)

While most FcγRs (FcγRIIa, IIb, and IIIa/b) have low affinity to monomeric Fc, ICs contain a higher density of Fcs that can cross link and activate FcγRs. To assess IC formation between tobevibart and HBsAg, we used transmission electron microscopy (TEM) to examine a range of concentrations of tobevibart and HBsAg SVPs by ammonium molybdate negative staining. In the absence of mAb, only individually dispersed, spherical HBsAg SVPs were observed. ICs were identified in the presence of tobevibart as aggregated clusters of HBsAg SVPs. At 1,500 IU/mL HBsAg, the peak size of ICs occurred with 5 µg/mL of tobevibart. IC size decreased at the highest tobevibart concentration of 500 µg/mL (Fig. 1). At 4,500 IU/mL HBsAg, higher concentrations of tobevibart (∼50 µg/mL) were required to form the largest ICs. Results were consistent in a total of five similar experiments done at slightly different temperatures, concentrations, or incubation times. These data demonstrated that tobevibart forms ICs with HBsAg and that the ratio of tobevibart and HBsAg influences IC size.

**Figure 1:**
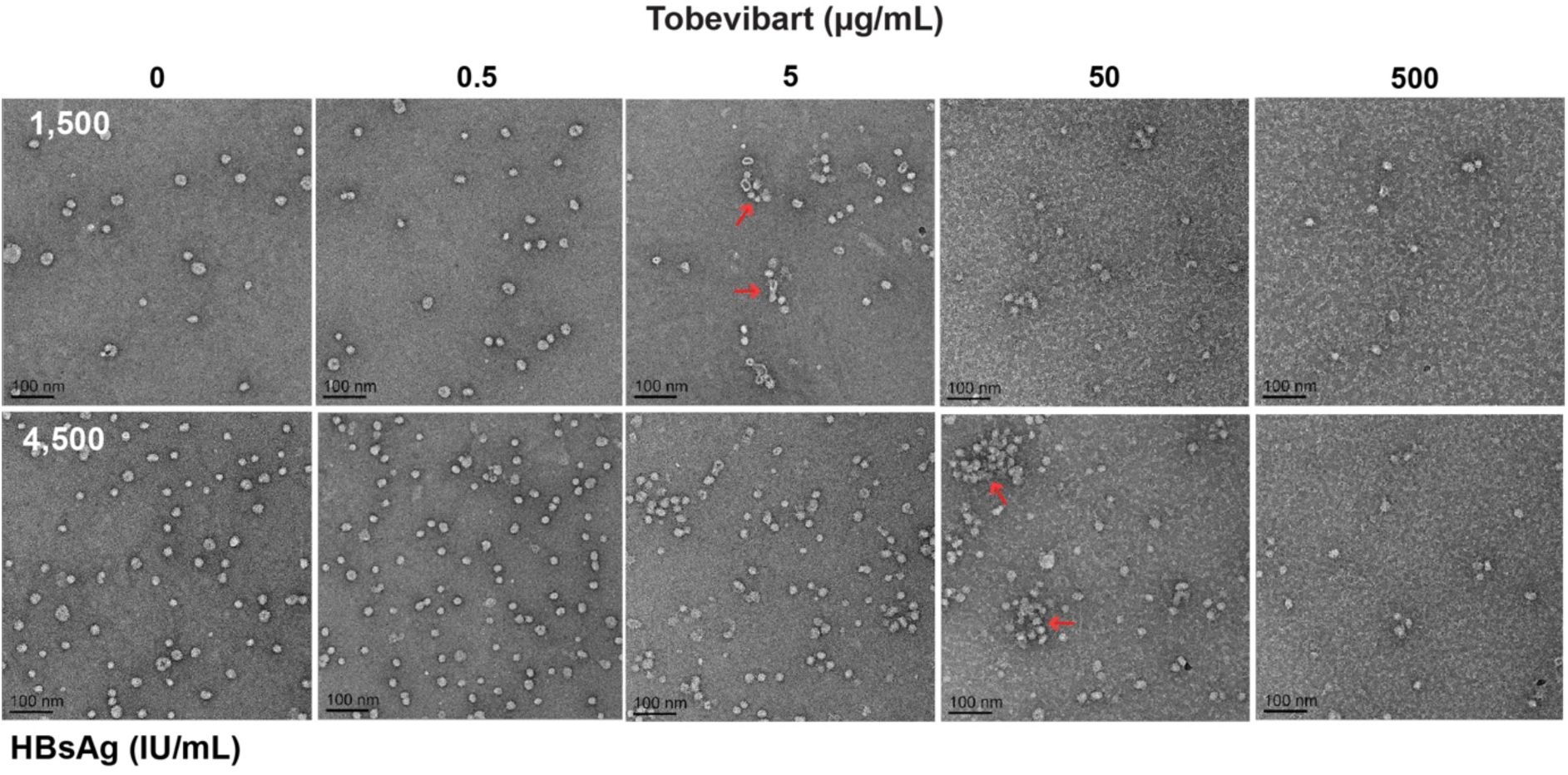
Immune complex (IC) formation of tobevibart with HBsAg. Electron microscopy images of HBsAg subviral particles (SVPs) pre-incubated with tobevibart at different concentrations. Single HBsAg SVPs (round vesicles) and tobevibart-HBsAg ICs that appear as aggregates (red arrows point to largest ICs) were visualized by negative staining. HBsAg concentrations were 1,500 or 4,500 IU/mL, and tobevibart was 0, 0.5, 5, 50, or 500 µg/mL. One representative image per sample.

### The engineered GAALIE Fc of tobevibart increased Fc**γ**R signaling

We next examined the efficiency of ICs at different ratios of tobevibart and HBsAg derived from PLC/PRF/5 hepatoma cells for inducing human FcγR signaling in a Jurkat cell NFAT luciferase reporter assay. Peak activation of FcγRs was induced at an optimal ratio of 1,000 IU/mL HBsAg and 20 µg/mL of tobevibart for FcγRIIa and 3,330 IU/mL HBsAg and 1.6-6.2 µg/mL of tobevibart for FcγRIIIa signaling (Fig. 2A). Activation of FcγRIIa was confirmed using HBsAg derived from patient serum (Suppl. Fig. S1). These data demonstrated that optimal concentrations of HBsAg and tobevibart are needed to efficiently trigger FcγRs (Fig. 2B).

**Figure 2:**
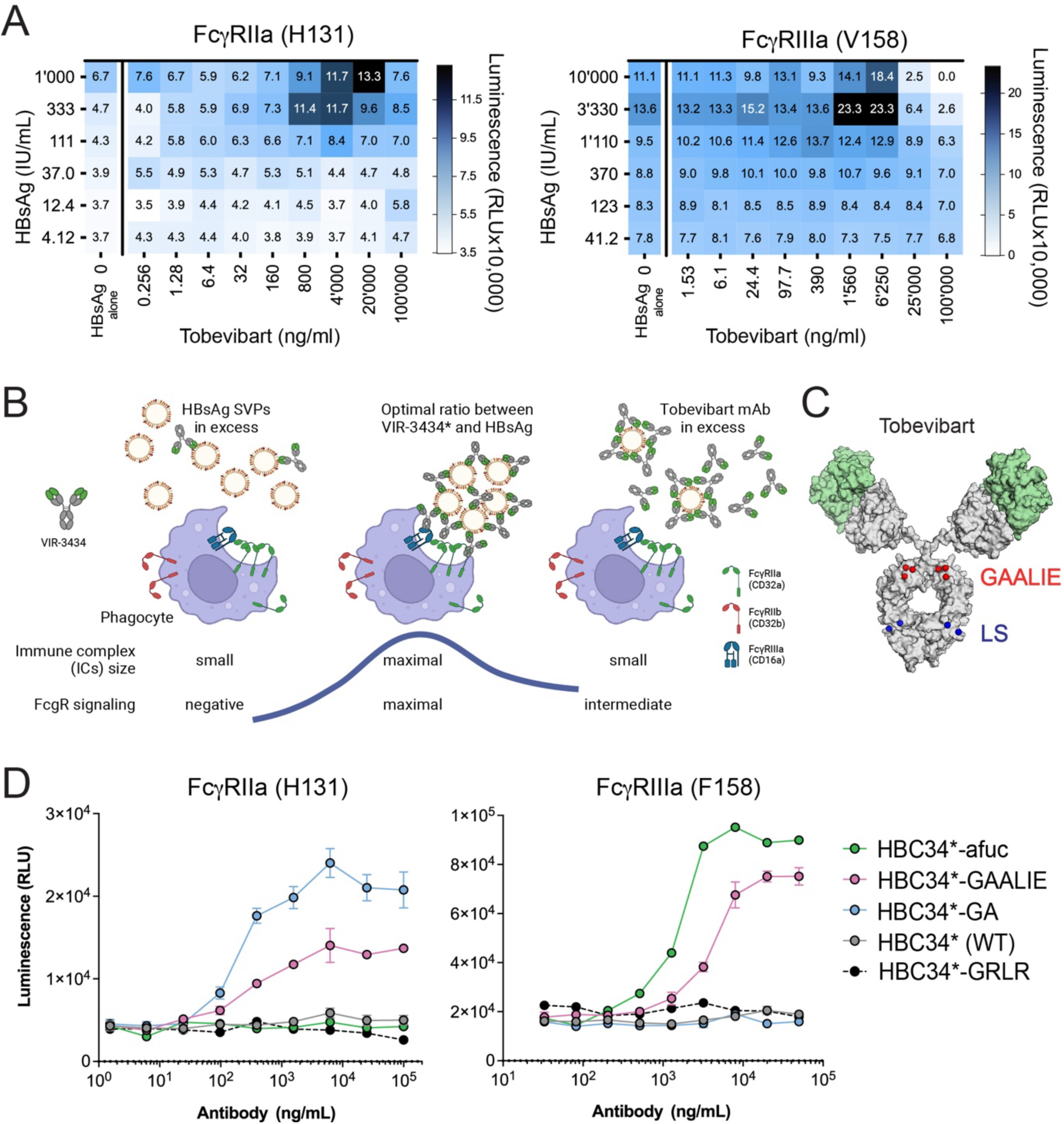
FcγRIIa and IIIa signaling induced via tobevibart / HBC34*-GAALIE. **(A)** Matrix of HBsAg (from PLC/PRF/5 cells) and tobevibart concentrations inducing FcγRIIa H131 (**left**) or FcγRIIIa V158 (**right**) signaling in Promega reporter cells. **(B)** Illustrated effect of tobevibart and HBsAg ratios influencing IC size, FcγR signaling, and phagocyte activation. High HBsAg and low mAb concentrations generate small or no ICs and no FcγR signal (**left**). Intermediate concentrations of HBsAg and mAb generate a maximal IC size and FcγR signal (**center**). Low HBsAg and high mAb concentrations generate small ICs and an intermediate FcγR signal (**right**). Created using Biorender. **(C)** Structural model of a tobevibart IgG molecule, illustrating the two Fab fragments (variable regions in green) and the Fc fragment, depicting the two heavy chains, which each include one GAALIE mutation (3 amino acids, red) and one LS mutation (2 amino acids, blue). **(D)** Signaling of FcγRIIa H131 (left) or FcγRIIIa F158 (right) induced by HBC34* Fc variants in ICs with 250 or 5,000 IU/mL recombinant HBsAg, respectively. Representative results of 2 independent experiments per FcγR. Error bars indicate standard deviation (SD) of two technical replicates.

A Fab (HBC34*) that differs by 3 amino acids in the light chain variable region from the clinical lead tobevibart was used as a surrogate in subsequent experiments. HBC34* with the same LS-GAALIE Fc as tobevibart was named HBC34*-GAALIE and maintained the same HBsAg binding, HBV neutralization, and FcγR activation profile as tobevibart (Suppl. Fig. S2). To better characterize the contribution of different FcγRs to the potential immunomodulatory effects of LS-GAALIE, we generated additional HBC34* Fc variants, all including the LS Fc mutation. The LS Fc (Fig. 2C) did not alter human FcγR binding but improved binding to the neonatal FcRn at acidic pH (Suppl. Fig. S2), rescuing mAbs from degradation more efficiently than a WT Fc [11]. The G236A (GA) Fc mutation increased binding only to FcγRIIa, the afucosylated Fc (afuc) of the N297 glycan increased binding only to FcγRIIIa, and the G326R-L328R (GRLR) Fc abrogated FcγR binding and served as negative control (Table 1, Suppl. Fig. S3)[15].

**Table 1:**
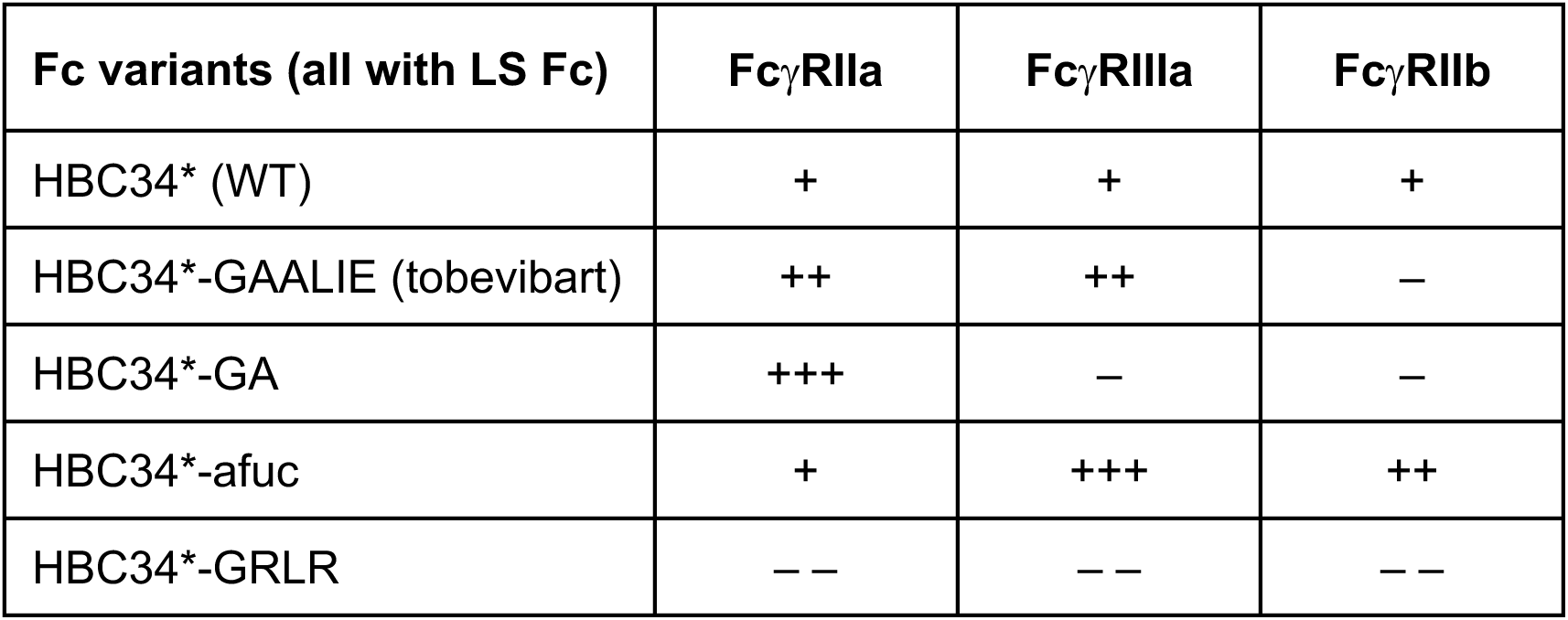
FcγR binding of HBC34* Fc variants.

| **Fc variants (all with LS Fc)** | **FcγRIIa** | **FcγRIIIa** | **FcγRIIb** |
| --- | --- | --- | --- |
| HBC34* (WT) | + | + | + |
| HBC34*-GAALIE (tobevibart) | ++ | ++ | – |
| HBC34*-GA | +++ | – | – |
| HBC34*-afuc | + | +++ | ++ |
| HBC34*-GRLR | – – | – – | – – |

When FcγR signaling of these Fc variants was examined in the presence of PLC-derived HBsAg, HBC34* with WT Fc or the GRLR negative control did not activate FcγRIIa and IIIa signaling (Fig. 2D). The GA and afuc Fc variants enabled binding and activation only of FcγRIIa or FcγRIIIa, respectively (Fig. 2D and Suppl. Fig. S3). In contrast, HBC34*-GAALIE (Fig. 2C) increased binding to activating FcγRs IIa and IIIa (as well as scavenger receptor FcγRIIIb) and decreased binding to the inhibitory FcγRIIb by surface plasmon resonance (Table 1 and Suppl. Fig. S3) and induced dose-dependent activation of both FcγRIIa and IIIa (Fig. 2D). Further, GAALIE abrogated binding to complement component C1q (Suppl. Fig. S2). Collectively, these data provide evidence that the GAALIE Fc enables a gain of function in the activation of, both, FcγRs IIa and IIIa, which has implications for enhanced immune cell engagement.

### HBC34*-GAALIE mediated Fc**γ**R-dependent binding of HBsAg ICs to immune cells *in vitro*

We next measured the binding of HBsAg ICs to immune cells *in vitro*, using whole blood samples from patients with CHB. The samples were incubated with HBC34*- GAALIE for 2 hours to form ICs with endogenous HBsAg and allow binding of ICs to FcγRs. Whole blood cells then were fixed and stained for analysis by flow cytometry using an anti-HBs mAb (HBi) that does not compete with HBC34* binding to HBsAg (Suppl. Fig. S4). Only in the presence (but not in the absence) of HBC34*-GAALIE was endogenous HBsAg detected via HBi staining on the surface of various immune cells (Fig. 3A and Suppl. Fig. S4 for gating). When HBC34*-GAALIE was added, HBsAg was primarily detected on the surface of neutrophils, CD16+ NK cells, and CD16+ non-classical (NC) monocytes, and, to lesser extent, on classical monocytes and cDCs (Fig. 3A). These data indicate that ICs formed by HBsAg and HBC34*- GAALIE can be captured by various immune cells.

**Figure 3:**
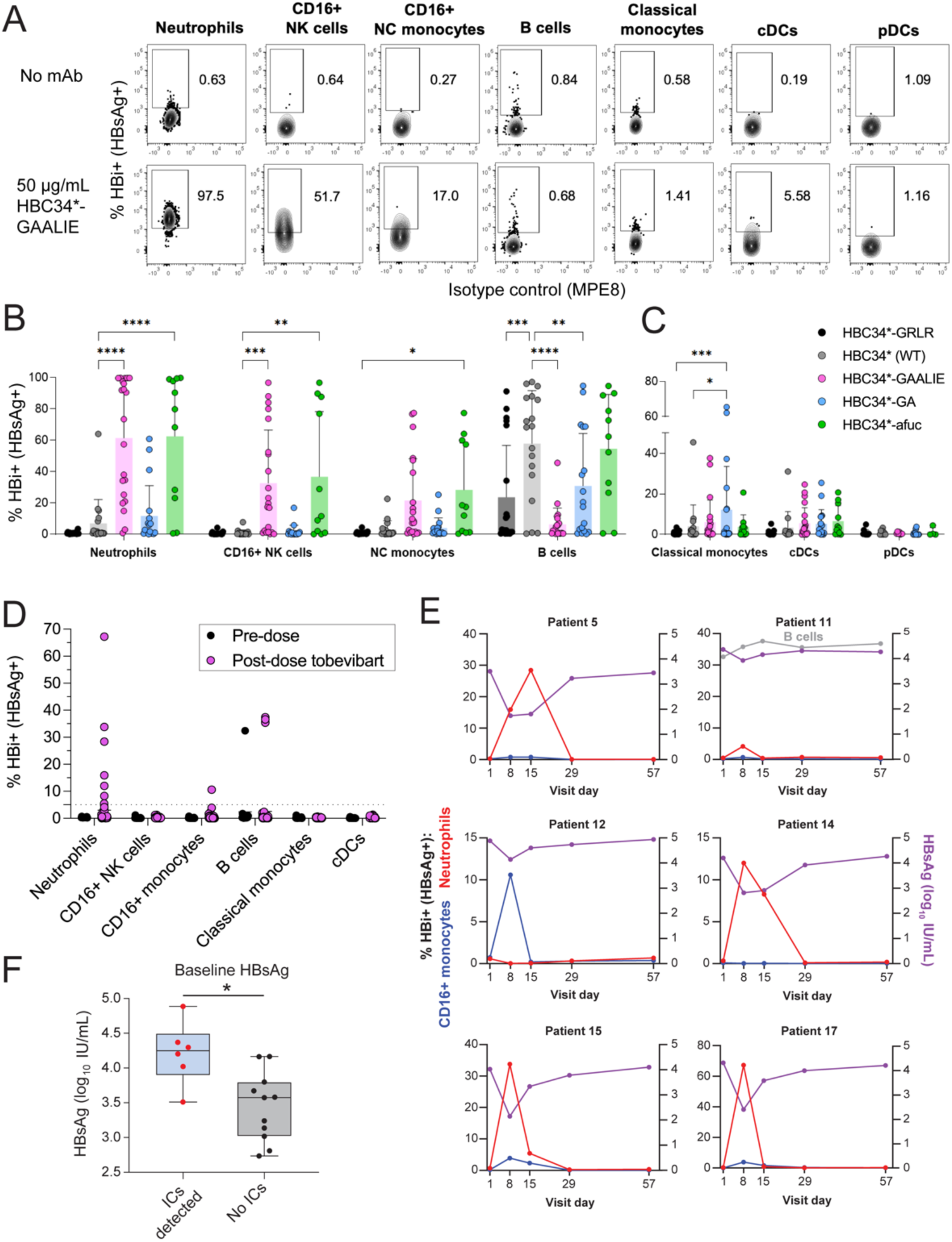
FcγR-mediated binding of HBsAg ICs with HBC34*-GAALIE or tobevibart to immune cells, *in vitro* and in a phase 1 clinical trial. **(A)** Representative flow cytometry plots of immune cells with or without addition of HBC34*-GAALIE *in vitro* into whole blood samples from patients with CHB. MAb HBi then was used to detect surface-bound HBsAg. **(B)** %HBi+ neutrophils, CD16+ NK cells, CD16+ NC monocytes, and B cells or **(C)** classical monocytes, cDCs and pDCs after adding 50 µg/mL of HBC34*-GAALIE or other Fc variants *in vitro* to HBV+ whole blood samples. Bars graph means ±SD and individual data points of independent experiments using n=11-21 blood donors. Statistics using 2-way ANOVA with Bonferroni correction for multiple comparisons. **(D)** Detection of %HBi+ HBsAg ICs on the surface of immune cells in whole blood samples collected directly in stabilizer from participants with CHB who received a single dose of 300 mg tobevibart in a phase 1 clinical trial. Each dot represents one of the collected time points (day 1, 8, 15, 29, and 57) pooled from all participants. A 5% HBi+ cutoff was applied to determine HBsAg positivity (dotted line). **(E)** Kinetics of HBsAg IC binding to neutrophils, CD16+ monocytes or B cells (left y-axis) and of HBsAg levels (IU/mL) in serum samples (right y-axis) obtained on the same day from patients with CHB post-dose of tobevibart. Kinetics are depicted for the 6 patients who had detectable HBi+ HBsAg ICs on any immune cell subset. **(F)** HBsAg levels (IU/mL) at baseline (pre-dose, day 1) of participants with detectable HBsAg ICs on immune cells or without ICs detected. Statistics using Mann-Whitney.

To assess the role of FcγRs, Fc variants of HBC34* were used (Table 1). HBi staining was low for the HBC34*-GRLR negative control, indicating that HBsAg binding in the presence of HBC34*-GAALIE was Fc dependent (Fig. 3B). B cells in a few samples stained positive for HBi after addition of HBC34*-GRLR (Fig. 3B), indicating some FcγR-independent binding of HBsAg ICs. Relative to HBC34* (WT Fc), addition of HBC34*-GAALIE resulted in significantly higher frequencies of HBsAg IC detection on FcγRIIIb^+^ neutrophils and CD16^+^ (FcγRIIIa^+^) NK cells, and, as a trend, on CD14^−^ CD16^+^ (FcγRIIIa^+^) NC monocytes (Fig. 3B). In addition, HBC34*-GAALIE or WT Fc mediated some binding of HBsAg to CD14^+^ CD16^−^ classical monocytes, cDCs, and plasmacytoid DCs (pDCs) (Fig. 3C). In contrast, significantly reduced frequencies of HBsAg IC-positive B cells were observed for HBC34*-GAALIE compared to WT Fc (Fig. 3B), consistent with reduced binding of the GAALIE Fc to the inhibitory FcγRIIb– –the only FcγR expressed on B cells (Suppl. Fig. S4). Weakening of the inhibitory interaction with FcγRIIb via HBC34*-GAALIE could potentially contribute to improved anti-HBs antibody responses.

We next dissected which FcγRs promoted the binding of HBsAg ICs to immune cells. Compared to WT Fc, addition of HBC34*-afuc (an Fc variant increasing binding only to FcγRIII) resulted in more HBsAg binding to FcγRIIIb+ neutrophils, CD16^+^ NC monocytes, and CD16^+^ NK cells, reaching similar levels as observed with the addition of HBC34*-GAALIE (Fig. 3B). This increased binding was also in line with the high expression of FcγRIII on these cell types (Suppl. Fig. S4). In contrast, addition of HBC34*-GA (increasing binding only to FcγRIIa) resulted in more HBsAg binding to classical monocytes and cDCs, reaching binding levels similar to HBC34*-GAALIE (Fig. 3C) and consistent with high expression of FcγRIIa on these immune cells (Suppl. Fig. S4). Consequently, HBC34*-GAALIE increased the capture of HBsAg ICs via FcγRIII by neutrophils, NC monocytes, and NK cells interaction and, to a lesser extent, via FcγRIIa by classical monocytes and cDCs. The binding of HBsAg ICs to a broad range of immune cells, which have potent phagocytic as well as anti-viral and antigen-presenting functions, may play a crucial role in reducing HBsAg levels in circulation.

### Tobevibart mediated HBsAg IC binding to neutrophils and CD16+ monocytes in a phase 1 clinical study

To assess translatability in vivo, we examined tobevibart-mediated delivery of endogenous HBsAg in ICs to immune cells in participants with CHB in a phase 1 clinical trial (VIR-3434-1002, ClinicalTrials.gov identifier NCT04423393). A single subcutaneous dose of 300 mg tobevibart was generally well tolerated and led to a rapid reduction in HBsAg in most participants with CHB [16]. Whole blood samples were collected directly into Cytodelics cell stabilizer pre-dose (Day 1) and post-dose of tobevibart on Days 8, 15, 29, and 57. For the detection of HBsAg ICs on the surface of immune cell subsets, samples were stained with HBi and cell markers for flow cytometric analysis (see Suppl. Tables S2 and S3 for lists of markers). Of the 18 total participants administered 300 mg, 17 had an analyzable Day 1 sample and were included in the IC evaluation. Following a single 300 mg dose of tobevibart, HBsAg ICs were detectable on circulating neutrophils (4 of 17 participants) and CD16+ monocytes (2 of 17 participants) (Fig. 3D, Suppl. Fig. S5). For participants with detectable ICs, HBsAg levels on neutrophils or CD16+ monocytes were generally highest at Day 8, when HBsAg reduction was the greatest (Fig. 3E). One of these participants (patient 11) also had HBsAg ICs detectable on B cells (Fig. 3D and E). Participants that carried detectable HBsAg ICs on the surface of immune cells had significantly higher baseline HBsAg, indicating that higher levels of HBsAg may have been required for IC detection (Fig. 3F). HBsAg IC binding to immune cells may have occurred before Day 8, but no samples were available on earlier time points. Overall, these data establish proof of concept that tobevibart forms ICs with endogenous HBsAg *in vivo* in patients with CHB and targets these complexes to immune cells, potentially leading to enhanced immune engagement.

### ICs of HBC34*-GAALIE and HBsAg activated human moDCs in vitro

Beyond immediate reduction of circulating HBsAg, the formation of ICs with HBC34*- GAALIE may induce activation of antigen-presenting cells, such as monocyte-derived DCs (moDCs). *In vitro* stimulation of primary human moDCs with HBC34*-GAALIE complexed with HBsAg from HBV+ patient serum increased the surface expression of activation marker CD83 (Fig. 4A,B). HBsAg ICs with HBC34*-GAALIE activated moDCs significantly more than ICs with HBC34* (WT Fc) at various HBsAg concentrations (Fig. 4A,B). In contrast, HBsAg ICs with HBC34*-GRLR did not activate moDCs, indicating that activation of moDCs depended on FcγRs (Fig. 4A,B). Activated moDCs secreted cytokines, such as IFNγ (Fig. 4C), IL-1β, IL-2, IL-6, and IL-10 (Suppl. Fig. S6) into culture supernatants. These results indicate that interaction of Fc with FcγRs was essential for the activation of moDCs and that the GAALIE Fc of HBC34* promoted significantly higher stimulation of moDCs than the WT Fc.

**Figure 4:**
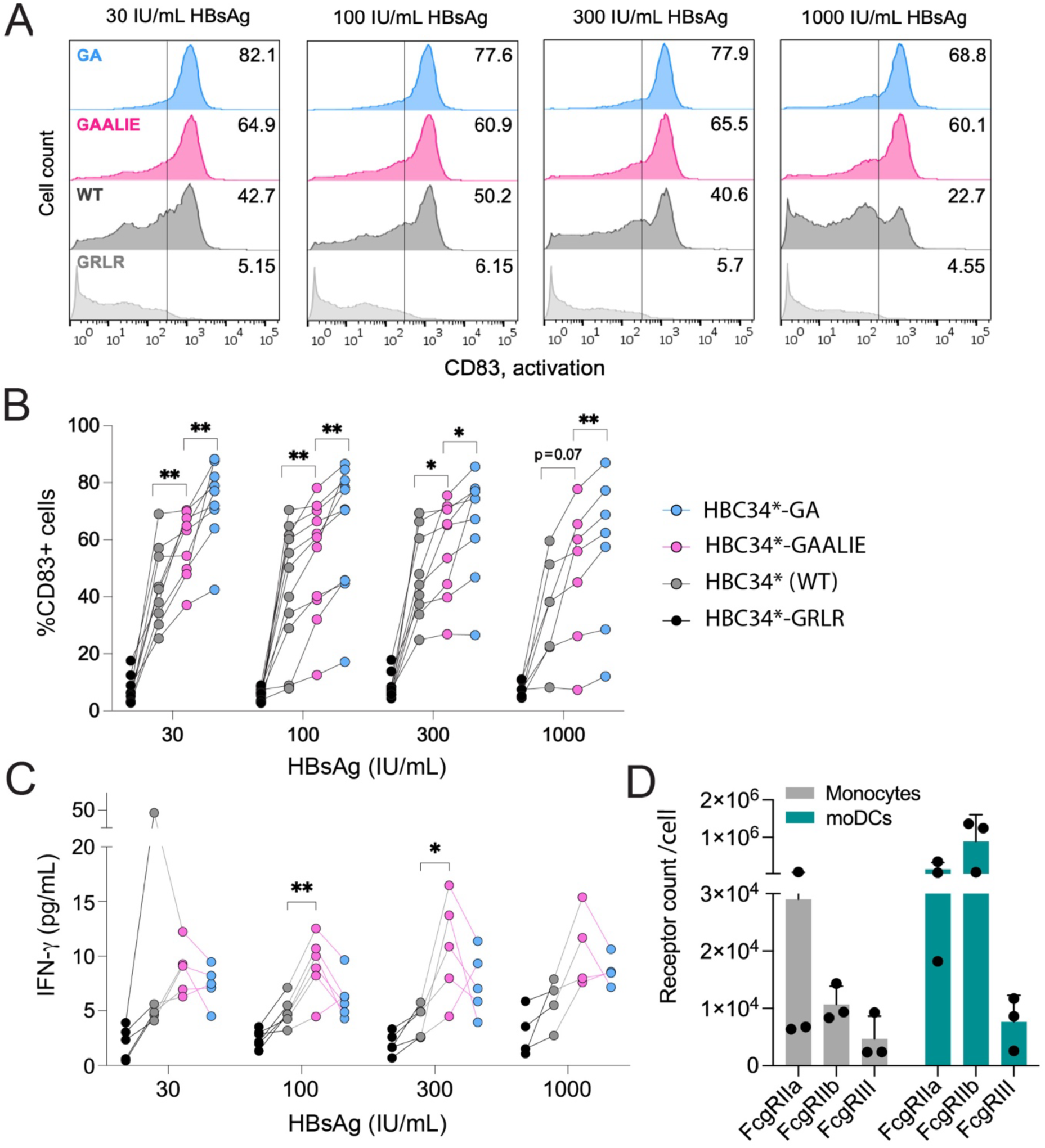
Human moDC activation via ICs of HBC34*-GAALIE and HBsAg. **(A)** Representative flow cytometry plots showing % CD83+ activated moDCs upon stimulation with 30-1,000 IU/mL HBsAg from HBV+ patient sera in complex with 50 μg/mL HBC34* Fc variants. **(B)** Pooled data showing % CD83+ moDCs of 5 independent experiments using 6 donors and 5 HBV+ patient sera described in A. Each line connects the same monocyte donor and HBV+ serum sample. The FcγRIIa genotype for 4 of 6 donors was known to be 131H/R. Statistical paired analysis used a 2-way ANOVA with Geisser-Greenhouse correction and Dunnett’s multiple comparison test. **(C)** Amount of IFN-γ secreted by moDCs stimulated as in A and B. Results from 3 monocyte donors and 5 HBV+ patient sera. Statistical paired analysis used a 2-way ANOVA with Geisser-Greenhouse correction and Dunnett’s multiple comparison test. **(D)** Expression of FcγRIIa, FcγRIIb, and FcγRIIIa/b on monocytes and moDCs (mean ±SD and individual data points of 3 independent experiments using 3 donors).

MoDCs express high levels of FcγRIIa (CD32a) and FcγRIIb (CD32b), and only a subset expresses FcγRIIIa (CD16a) (Fig. 4D). HBsAg ICs with HBC34*-GA induced significantly higher activation of moDCs compared to HBC34* (WT) or HBC34*- GAALIE (Fig. 4A and B), a result that is consistent with higher binding of the GA Fc to FcγRIIa (Suppl. Fig. 2). These results indicate that efficient FcγRIIa engagement is necessary for moDC activation. In contrast, HBC34*-GA does not increase binding to FcgRIII (Suppl. Fig. S3) and did not substantiate HBsAg IC binding to neutrophils or CD16+ monocytes (Fig. 3). Taken together, these data suggest that the GAALIE Fc of HBC34* ideally combines effective FcγRIII-mediated reduction of HBsAg (Fig. 3) with increased activation of moDCs via FcγRIIa (Fig. 4), which in turn may induce long-lasting HBsAg-specific T cell responses.

### HBC34*-GAALIE ICs in vitro activated human HBsAg-specific CD4+ memory T cells induced by HBV vaccination

To develop long-lasting adaptive immunity against HBV, activated DCs must present HBsAg-derived peptides to T cells. To evaluate this process *in vitro*, total CD4+ memory T cells isolated from HBV vaccinees were co-cultured with autologous moDCs that had been stimulated with ICs formed by HBsAg from HBV+ patient sera and HBC34* Fc variants (Fig. 5A). After 5 days, HBC34*-GAALIE or the WT Fc induced similar degrees of proliferation of HBsAg-specific CD4^+^ memory T cells as measured by CFSE staining and the expression of activation marker CD25 (Fig. 5B and C). These results suggest that potential effects of Fc engineering on enhancing T cell proliferation were not resolved in this assay beyond that stimulated by WT IgG1, likely due to the high variability when using primary human T cells. In contrast, when moDCs were stimulated with HBsAg in complex with HBC34*-GRLR (abrogated binding to FcγRs), CD4^+^ T cell proliferation and activation were significantly lower (Fig. 5B and C) and comparable to stimulation with HBsAg alone (Suppl. Fig. S7), confirming the Fc dependence of antibody enhanced T cell responses.

**Figure 5:**
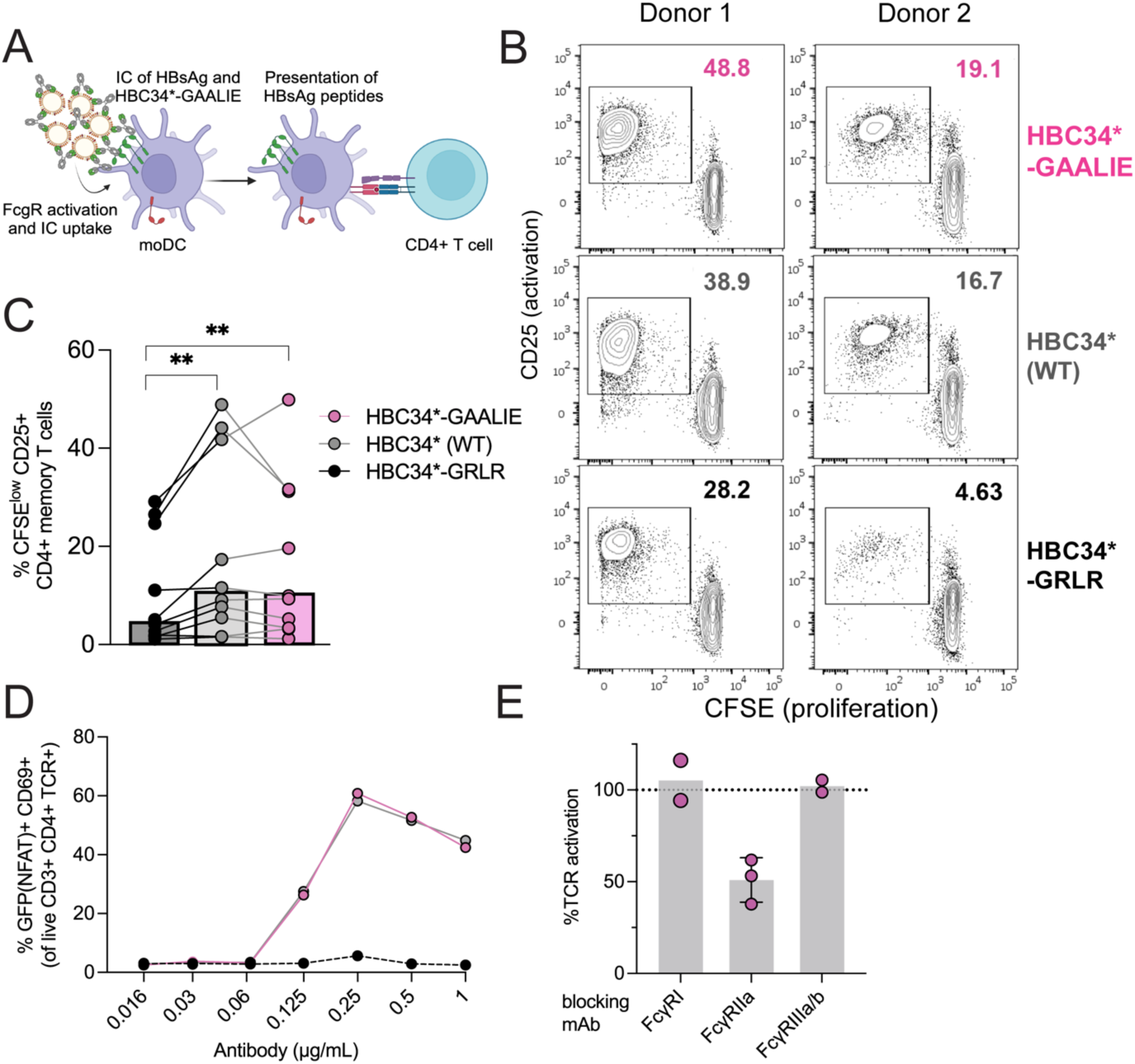
Reactivation of human, HBsAg-specific CD4+ memory T cells from HBV vaccinated donors. **(A)** Experimental setting of IC-mediated stimulation of moDCs, followed by co-culture with human CD4+ T cells. Image generated using Biorender. **(B)** CD25 expression and CFSE dilution of CD4+ memory T cells in co-culture with autologous moDCs stimulated with ICs of 100 IU/mL HBsAg from HBV+ patient serum and 50 μg/mL HBC34*- GAALIE or Fc variants (WT, GRLR) for two representative monocyte donors. **(C)** Summary graph showing median (bars) and individual data points for % CFSE low CD25+ activated CD4+ memory T cells from 5 independent experiments using PBMCs from 3 HBV vaccinees (FcγRIIa-131H/H, H/R, and R/R) and 100 IU/mL HBsAg from 3 HBV+ patient sera. Wilcoxon matched-pairs signed rank test. **(D)** % GFP+ CD69+ activated TCR-transgenic reporter cells specific for HBsAg in co-culture with HLA-matched moDCs stimulated with HBsAg from PLC cells at 1,000 IU/mL in ICs with HBC34*- GAALIE or Fc variants (WT, GRLR). Representative data of six independent experiments using two monocyte donors. **(E)** Experiment as in **(D)** pre-treating moDCs with 1 µg/mL of anti-FcγRs blocking mAbs prior to stimulation with HBsAg IC and HBC34*-GAALIE. Bar graph shows means ±SD, normalized to the non-blocked control. Individual data points of three monocyte donors and 3 independent experiments.

These results were recapitulated in co-cultures of HBsAg IC-stimulated moDCs with Jurkat reporter cells transgenic for a T cell receptor (TCR) that recognizes an HBsAg peptide presented on HLA-DR. TCR triggering in this model leads to NFAT-mediated GFP expression. A significant increase in %GFP+ CD69+ activated Jurkat cells was observed when moDCs were stimulated with HBsAg ICs including HBC34*- GAALIE or the WT Fc compared to stimulation with HBsAg alone or HBC34*-GRLR (Fig. 5D, Suppl. Fig. S7). Moreover, blocking of FcγRIIa (but not FcγRI or III) on moDCs resulted in a ∼50% decrease in TCR activation, following stimulation with ICs of HBsAg and HBC34*-GAALIE (Fig. 5E). Thus, the HBC34*-GAALIE Fc interaction with FcγRIIa enabled moDCs to present HBsAg-derived peptides and stimulate CD4+ T cell responses−an interaction that may potentially contribute to long-lasting immunity and functional cure of CHB.

### HBC34*-GAALIE ICs in vitro activate polyclonal CD4+ memory T cells of HBV-vaccinated mice more efficiently than ICs of the mAb and the WT Fc

To evaluate T cell activation in an orthogonal system, we examined the *ex vivo* reactivation of HBsAg-specific CD4+ memory T cells from HBV-vaccinated C57BL/6 mice after co-culture with bone marrow-derived DCs (BMDCs) from HuFcγR syngeneic mice [13]. This model allows investigation of human Fc variants, such as GAALIE, which specifically modulate binding to human but not mouse FcγRs. C57BL/6 mice were vaccinated with the Engerix B20 HBV vaccine and boosted after two weeks. Two months after the booster, total CD44+ CD4+ memory T cells were sorted from lymph nodes and spleens by flow cytometry and labelled with CFSE. In parallel, BMDCs were generated from HuFcγR mice and stimulated overnight with ICs of HBsAg from PLC hepatoma cells and HBC34*-GAALIE, WT or GRLR. Next, pulsed BMDCs were co-cultured for 6 days with the CFSE-labelled CD4+ memory T cells and their proliferation was assessed by flow cytometry (Fig. 6A, Suppl. Fig. S8). HuFcγR-BMDCs induced a significantly higher proliferation of HBsAg-specific CD4+ memory T cells when stimulated with HBC34*-GAALIE ICs across a broad range of mAb concentrations compared to ICs formed with HBC34* with WT Fc (Fig. 6B-C, Suppl. Fig. S8). In contrast, the level of proliferation of CD4^+^ memory T cells in cultures with the HBC34*-GRLR negative control was comparable to that of unstimulated, HBsAg-alone, or mAb-alone controls (Fig. 6B and D). Collectively, these results indicate that the GAALIE Fc can efficiently mediate antigen presentation and expansion of HBsAg-specific CD4+ memory T cells, supporting the concept that Fc engineering can amplify adaptive T cell immunity against HBV.

**Figure 6:**
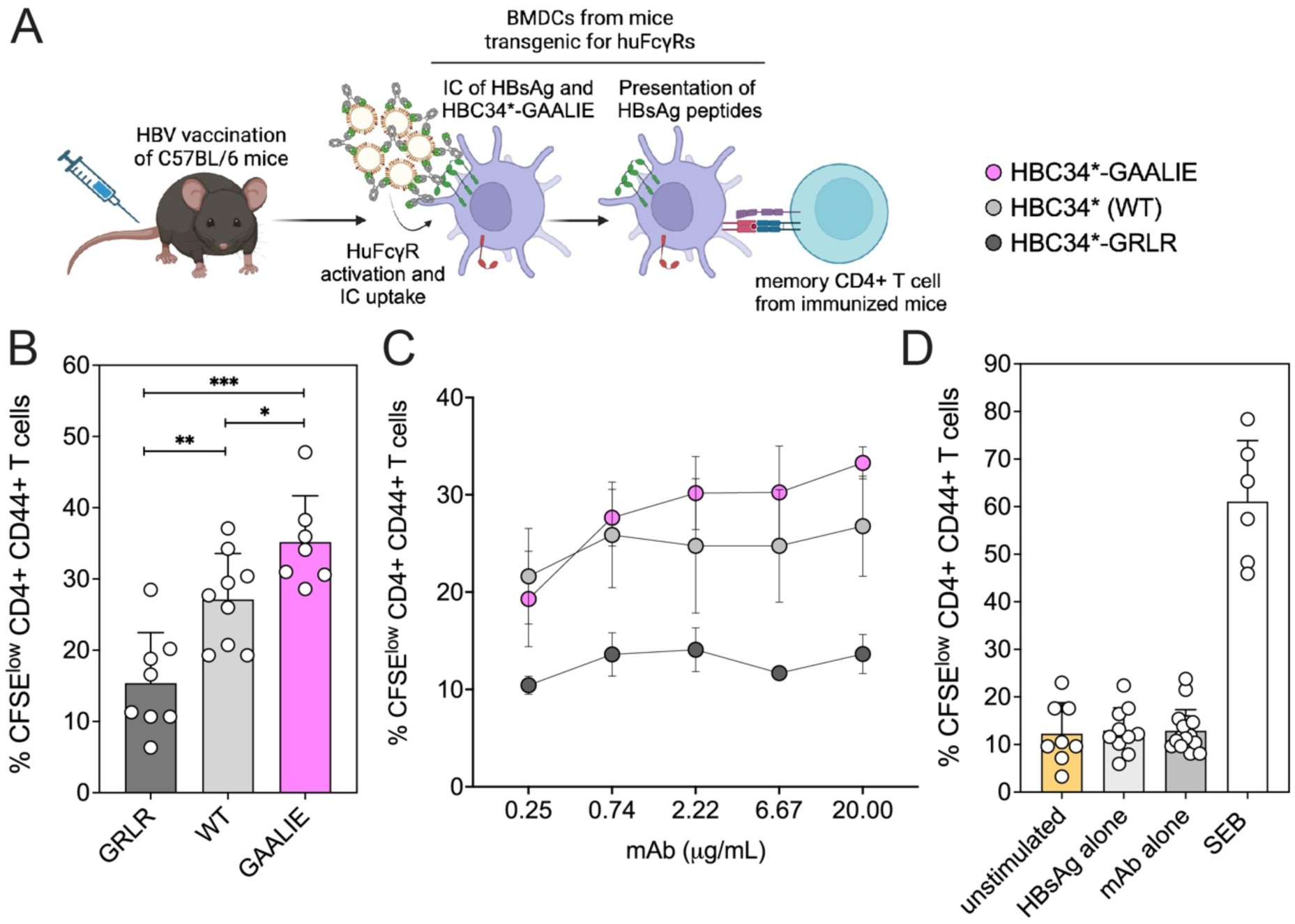
Reactivation of murine CD4+ memory T cells *in vitro*, using cells from HBV-vaccinated and HuFcγR mice. **(A)** Experimental setting of HBV vaccination of mice, followed by *in vitro* co-culture of BM-DCs stimulated with HBsAg ICs and memory CD4+ T cells from immunized mice. Image generated using Biorender. **(B)** Pooled data (means ±SD and individual data points) from 9 independent experiments of % CFSE low CD44+ CD4+ memory T cells re-stimulated *in vitro* with ICs containing 1,000 IU/mL HBsAg and 20 µg/mL HBC34* Fc variants. **(C)** Curves showing % CFSE low CD44+ CD4+ memory T cells re-stimulated *in vitro* with ICs containing 1,000 IU/mL HBsAg and titrated HBC34* Fc variants. **(D)** Pooled data from control cultures containing HBsAg alone (1,000 IU/mL), mAb alone (20 µg/mL), SEB (1 µg/mL) or medium only (unstimulated). Statistical analyses used a Mann-Whitney nonparametric t-test.

## Discussion

New treatments for CHB aim to reverse immune exhaustion associated with prolonged infection and HBsAg tolerogenic effects and to induce functional cure by eliciting or expanding HBV-specific T and B cell responses. Immunotherapies comprised of mAbs to HBsAg that have the dual functions of Fab-mediated virus neutralization and enhanced Fc-mediated effector functions represent a promising approach towards achieving this goal. This study extends the characterization of Fab-mediated neutralization of HBV and HDV by tobevibart [6], adding evidence about how the engineered GAALIE Fc may function uniquely to enhance its therapeutic potential. Specifically, tobevibart mediated efficient capture of HBsAg by FcγRs on human immune cells−a mechanism that may enable phagocytosis and removal of HBsAg from the circulation. Capture of endogenous HBsAg by immune cells also occurred in patients with CHB dosed with tobevibart in a phase 1 clinical study. Further, we used several approaches to establish that ICs of tobevibart and HBsAg activated moDCs, enabling antigen presentation and the activation of HBsAg-specific memory T cells.

Previously we showed that tobevibart neutralized HBV and HDV *in vitro*, potentially by capturing virions into ICs [6]. Importantly, tobevibart was also highly effective in reducing HBV and HDV viremia and circulating HBsAg *in vivo* in a human liver-chimeric mouse model [6]. A recent study confirmed the value of this mouse model to monitor neutralizing mAb activity against dual infection [17]. Beyond Fab-mediated neutralization, the rapid reduction of circulating HBsAg by tobevibart [6] may be mediated by Fc-dependent mechanisms that are enhanced by Fc engineering. While HBsAg alone can be taken up at low levels by human B cells and monocytes, HBsAg complexed with anti-HBs antibodies containing WT Fcs can be taken up at higher levels via FcγRs by B cells, monocytes, DCs and neutrophils [8]. Other mAbs with WT Fcs mediated HBsAg uptake by neutrophils, macrophages, and Kupffer cells in HBV-transgenic mice [7]. Fc-mediated effector functions were critical for phagocytosis and HBsAg removal, since these effects were abrogated by a mutation that abolish FcγR binding [7, 18].

Fc engineering that increases binding to mouse FcγRs II and III (human FcγRIIb and IIa equivalents) improved neutrophil and monocyte-mediated clearance of HBsAg in AAV-HBV mice, along with augmenting phagocytosis *in vitro* [19]. Here we report that tobevibart / HBC34*-GAALIE formed ICs with endogenous HBsAg and mediated IC binding *in vitro* via FcγRs to immune cells recovered from blood of patients with CHB. Specifically, the engineered GAALIE Fc of HBC34*-GAALIE enhanced HBsAg binding to neutrophils, CD16+ NC monocytes, and NK cells via engagement of FcγRIII binding. To a lesser extent, tobevibart / HBC34*-GAALIE also mediated binding of HBsAg to classical monocytes and cDCs, likely via FcγRIIa. *In vivo* proof of concept was provided by direct detection of HBsAg IC binding to immune cells in a phase 1 clinical trial of patients with CHB receiving a single dose of 300 mg tobevibart. ICs of tobevibart and HBsAg were present on the surface of circulating neutrophils and CD16+ monocytes primarily on Day 8, when the reduction of circulating HBsAg levels was most pronounced. We speculate that the binding of HBsAg ICs to immune cells via FcγRs may be the first step in mediating phagocytosis and enable the clinically observed reduction of HBsAg levels following a single dose of tobevibart.

A recent study using an HBsAg mutation that abrogated secretion from hepatocytes in an AAV-HBV mouse model showed that eliminating HBsAg in circulation regenerated anti-HBs antibodies, but not T cell responses, following therapeutic vaccination [20]. Thus, short-term clearance of circulating HBsAg via phagocytosis alone may not be sufficient to reverse tolerance and achieve functional cure, which likely requires adaptive T cell immunity. In keeping with this hypothesis, resolution of acute HBV infection is associated with the induction of HBV-specific T cells [2]. Uptake of antigen and activation of antigen-presenting cells are the first critical steps towards eliciting or expanding antigen-specific T cells. Although altered frequencies and impaired functionality of cDCs and pDCs have been observed in some patients with CHB [21], depots of HBsAg were detected in monocytes from patients with CHB that did not impair their functionality [22]. Once differentiated to moDCs *in vitro*, these cells were able to present antigen and stimulate autologous HBV-specific CD8+ and CD4+ T cells [22]. Similarly, moDCs generated *in vitro* from patients with CHB were able to stimulate autologous T cells, when pulsed with recombinant HBV antigens [23]. In a clinical trial, re-infusion of autologous HBsAg-pulsed moDCs and T cells that were activated *in vitro* led to a decrease in HBV viral load and HBV DNA loss in some patients with CHB [24]. We found that tobevibart / HBC34*-GAALIE in complex with HBsAg effectively stimulated both FcγRIIa and IIIa signaling and activated moDCs *in vitro*, whereas an Fc variant (GRLR) with abrogated binding to FcγRs did not. This indicates the first critical steps in immune engagement are mediated by the engineered Fc. Further, these activated moDCs successfully presented HBsAg-derived peptides and activated CD4+ memory T cells from human HBV vaccinees, as well as HBsAg-specific TCR-transgenic CD4+ reporter T cells. These observations *in vitro*, together with our detection of ICs formed by tobevibart and endogenous HBsAg on circulating immune cells *in vivo*, suggest that this Fc-mediated mechanism may support the induction of HBsAg-specific T cells in CHB patients.

The efficacy of mAbs for treating CHB is likely limited not only by the exhausted state of HBsAg-specific memory T cells but also by suboptimal ratios of antibodies:HBsAg, leading to formation of small ICs that have low avidity for FcγRs. To compensate for this clinical circumstance, Fc engineering may play a pivotal role in enhancing antibody-dependent effector functions and extending the range of antibody:HBsAg ratios that can induce efficient FcγR engagement. In a study of prophylaxis against influenza virus, the GAALIE Fc mutation of an anti-HA stem mAb increased maturation of cDCs, augmented priming of naïve CD4^+^ and CD8^+^ T cells, and improved efficacy by 5-fold over the mAb with WT Fc in a mouse model expressing human FcγRs [15]. Our study indicates that human moDCs become significantly more activated when stimulated with tobevibart / HBC34*-GAALIE and HBsAg ICs compared to ICs formed by HBC34* with WT Fc. Additionally, HBC34*-GA, which highly increases binding only to FcγRIIa, efficiently activated moDCs, indicating a key role for FcγRIIa in this activation. Most likely, the decreased binding of the GAALIE Fc to the inhibitory FcγRIIb was also important to enable activation of moDCs, which express high levels of FcγRIIb. Likewise, CD4+ memory T cells from HBV-vaccinated mice were significantly more activated when re-stimulated *ex vivo* with HBC34*-GAALIE HBsAg ICs as compared to ICs formed with HBC34* with WT Fc. Consequently, Fc engineering leads to higher DC activation by ICs and may generate more potent stimulation of HBsAg-specific T cell immunity over the course of mAb treatment.

The capacity of antibodies to overcome T cell exhaustion was suggested by a study in mice tolerized to HBV antigens in which four doses of anti-HBs mAb (E6F6, containing a WT Fc) increased the frequency of HBsAg-specific CD8+ T cells [7]. However, a mAb that was Fc engineered to increase binding to mouse FcγRs led to more functional CD4^+^ and CD8^+^ T cells in the liver as well as HBsAg-specific B cells in the lymph nodes of tolerized AAV-HBV mice [19]. In the same model, treatment with anti-HBs mAb Ab-H that reduced HBsAg levels, restored the ability of CD4^+^ T cells or B cells to respond to subsequent therapeutic vaccination with recombinant HBsAg and of CD8^+^ T cells when the vaccination was further adjuvanted with CpG [10]. Similarly, prior reduction of HBsAg and HBV gene expression via siRNA increased the efficacy of therapeutic vaccination against chronic infection with HBV in mice [10, 20, 25]. Overall, mAb Fc engineering that provides specific activation of FcγR-expressing antigen presenting cells by ICs formed with HBsAg in patients with CHB in combination with other approaches to reduce HBsAg levels is likely the most promising strategy to achieve functional cure.

Future work of interest will be to assess the capacity of tobevibart to restimulate anergic or prime naïve CD4^+^ T cells or of CD8^+^ T cells, which requires cross-presentation of extracellular HBsAg on MHC class-I. Liver sinusoidal endothelial cells (LSECs) are key scavenger cells that efficiently clear small ICs [26, 27] via FcγRIIb-mediated endocytosis. LSECs cross-presented soluble antigen but not antigen captured via FcγRIIb in ICs [28], potentially hindering antibody-mediated induction of CD8+ T cell responses, in line with the tolerogenic liver environment. Thus, the decreased FcγRIIb binding of tobevibart GAALIE Fc may limit uptake of HBsAg-containing ICs into LSECs and, instead, make ICs available to other antigen-presenting cells within the liver or peripheral lymphoid organs.

In conclusion, these findings suggest that tobevibart has the potential to not only neutralize HBV and HDV infection but to decrease HBsAg levels and enable the induction of long-lasting adaptive immune responses against HBV necessary to achieve functional cure. These attributes are uniquely mediated via the engineered GAALIE Fc that combines increased binding to FcgRs IIa and III, while decreasing binding to inhibitory FcgRIIb. Based on its multiple modes of action, tobevibart provides a novel therapeutic with functions complementary to those of other therapies and is currently under clinical assessment for therapy of both CHB and CHD.

## List of Abbreviations

AAV: Adeno-associated virus
BMDCs: Bone marrow-derived dendritic cells
cDCs: Classical dendritic cells
CFSE: Carboxyfluorescein succinimidyl ester
CHB: Chronic hepatitis B
CHD: Chronic hepatitis Delta
FACS: Fluorescence activated cell sorter
FcγRs: Fc gamma receptors
FcRn: Neonatal Fc receptor
GFP: Green fluorescent protein
HBIG: hepatitis B immunoglobulin
HBsAg: Hepatitis B surface antigen
HBV: Hepatitis B virus
HDV: Hepatitis Delta virus
HuFcγR: Humanized FcγR
IC: Immune complex
LSEC: Liver sinusoidal endothelial cells
mAb: Monoclonal antibody
moDCs: Monocyte-derived dendritic cells
NC: Non-classical
NK cells: Natural killer cells
PBMC: Peripheral blood mononuclear cell
pDCs: Plasmacytoid dendritic cells
SD: Standard deviation
SEB: Staphylococcal enterotoxin B
SEC: Size exclusion chromatography
SVP: Subviral particle
TCR: T cell receptor
TEM: Transmission electron microscopy
WT: Wild-type

## Material & Methods

For details regarding the materials and methods used, please refer to the supplemental information and CTAT table.

## Acknowledgements

We thank Alan Korman, Adam Gehring, and Saiful Islam for advice and generating Jurkat TCR reporter cells. We thank Hannah Kaiser for running HBV neutralization assays, Josh R. Dillen for running preliminary SPR experiments, and Kevin Hauser for generating a mAb structural model.

## Data availability statement

All source data that support the findings of this study are available from the corresponding authors upon reasonable request.

## Competing interests

LV, RW, RM, BG, ES, SVG, LER, YPC, JdI, AM, KET, SD, JME, LW, NC, JD, NS, AP, LS, DCloutier, GS, CHT, FAL, CHD, FB, AL, AA, DCorti, MAS are or were employees of Vir Biotechnology and may hold shares in Vir Biotechnology. LER, NC, GS, FAL, DC and MAS are listed as inventors on patent applications, which disclose the subject matter described in this manuscript. KA, MFY, HW, and EG served as advisors or received grant support from various industry partners. The remaining authors declare no conflict of interest.

## Financial support

David Belnap received funding from Vir Biotechnology through an agreement with University of Utah, related to the work described in this paper. The tobevibart phase 1 clinical study received editorial support by Lumanity Scientific Inc., which was funded by Vir Biotechnology.

## Author contributions

Conceived study: LV, LER, LS, AL, CHT, FB, CHD, AA, DCorti, M.A.S. Performed experiments *in vitro*: LV, RW, BG, ES, JD, NS, JME, AP. Performed experiments *in vivo*: RM. Performed EM imaging: DMB. Conducted the phase 1 tobevibart clinical study and data analysis: RW, SVG, LW, Y-PC, JdI, AM, KET, DCloutier, AA, KA, MFY, HW, EG. Manuscript writing: LV and MAS. Supervision and manuscript editing: LER, NC, FAL, CHD, GS, AA, DCorti and MAS.

## Notes

### Clinical Trial

NCT04423393

### Funding Statement

This clinical trial was designed, conducted and funded by Vir Biotechnology, Inc. as sponsor. David Belnap received funding from Vir Biotechnology through an agreement with University of Utah, related to the work described in this paper. The tobevibart phase 1 clinical study received editorial support by Lumanity Scientific Inc., which was funded by Vir Biotechnology.

### Author Declarations

(A) For the phase 1 clinical trial (VIR-3434-1002, ClinicalTrials.gov, NCT04423393) Approval from the local Institutional Review Board or Independent Ethics Committee was obtained and informed consent was obtained from all participants prior to their participation in the study. These local review boards or committees were of the Medical Faculty of the University of Duisburg-Essen, Germany; London City & East Research Ethics Committee, Bristol, United Kingdom; University Hospital Birmingham, United Kingdom; Queen Mary Hospital, Hong Kong; University of Hong Kong, Hong Kong; Ethics Committee at Medical Center of Arensia Exploratory Medicine Limited Liability Company, Kyiv, Ukraine; Asan Medical Center, Seoul, South Korea; Gangnam Severance Hospital Yonsei University, South Korea; Korea University Anam Hospital, South Korea; Pusan National University Hospital, South Korea; Seoul National University Hospital, South Korea; Health and Disabilities Ethics Committee, Ministry of Health, Wellington, New Zealand; Romania Academy of Sciences, Bucharest, Romania; Singapore Health Services, Singapore. (B) For human samples obtained outside the above study for research in vitro, Whole blood, PBMCs, sera or plasma samples were obtained from human subjects under study protocols approved by the local Institutional Review Boards (Ethics Committees of the Canton Ticino, Switzerland or IRB of the University of California San Francisco or Advarra IRB for Quest Diagnostics, USA). All donors provided written informed consent for the use of blood and blood components.

## References

[1] WHO. Global hepatitis report 2024: action for access in low- and middle-income countries. online; 2024.

[2] Iannacone M, Guidotti LG. Immunobiology and pathogenesis of hepatitis B virus infection. Nat Rev Immunol 2022;22:19–32.

[3] Ye B, Liu X, Li X, Kong H, Tian L, Chen Y. T-cell exhaustion in chronic hepatitis B infection: current knowledge and clinical significance. Cell Death Dis 2015;6:e1694.

[4] Wong GLH, Gane E, Lok ASF. How to achieve functional cure of HBV: Stopping NUCs, adding interferon or new drug development? Journal of hepatology 2022;76:1249–1262.

[5] Corti D, Benigni F, Shouval D. Viral envelope-specific antibodies in chronic hepatitis B virus infection. Current opinion in virology 2018;30:48–57.

[6] Lempp FA, Volz T, Cameroni E, Benigni F, Zhou J, Rosen LE, et al. Potent broadly neutralizing antibody VIR-3434 controls hepatitis B and D virus infection and reduces HBsAg in humanized mice. Journal of hepatology 2023;79:1129–1138.

[7] Zhang TY, Yuan Q, Zhao JH, Zhang YL, Yuan LZ, Lan Y, et al. Prolonged suppression of HBV in mice by a novel antibody that targets a unique epitope on hepatitis B surface antigen. Gut 2016;65:658–671.

[8] Tharinger H, Rebbapragada I, Samuel D, Novikov N, Nguyen MH, Jordan R, et al. Antibody-dependent and antibody-independent uptake of HBsAg across human leucocyte subsets is similar between individuals with chronic hepatitis B virus infection and healthy donors. J Viral Hepat 2017;24:506–513.

[9] Celis E, Chang TW. Antibodies to hepatitis B surface antigen potentiate the response of human T lymphocyte clones to the same antigen. Science 1984;224:297–299.

[10] Zhu D, Liu L, Yang D, Fu S, Bian Y, Sun Z, et al. Clearing Persistent Extracellular Antigen of Hepatitis B Virus: An Immunomodulatory Strategy To Reverse Tolerance for an Effective Therapeutic Vaccination. J Immunol 2016;196:3079–3087.

[11] Gaudinski MR, Coates EE, Houser KV, Chen GL, Yamshchikov G, Saunders JG, et al. Safety and pharmacokinetics of the Fc-modified HIV-1 human monoclonal antibody VRC01LS: A Phase 1 open-label clinical trial in healthy adults. PLoS Med 2018;15:e1002493.

[12] Weitzenfeld P, Bournazos S, Ravetch JV. Antibodies targeting sialyl Lewis A mediate tumor clearance through distinct effector pathways. The Journal of clinical investigation 2019;129:3952–3962.

[13] Smith P, DiLillo DJ, Bournazos S, Li F, Ravetch JV. Mouse model recapitulating human Fcgamma receptor structural and functional diversity. Proceedings of the National Academy of Sciences of the United States of America 2012;109:6181–6186.

[14] Yamin R, Jones AT, Hoffmann HH, Schafer A, Kao KS, Francis RL, et al. Fc-engineered antibody therapeutics with improved anti-SARS-CoV-2 efficacy. Nature 2021;599:465–470.

[15] Bournazos S, Corti D, Virgin HW, Ravetch JV. Fc-optimized antibodies elicit CD8 immunity to viral respiratory infection. Nature 2020;588:485–490.

[16] Agarwal KY, M. F.; Wedemeyer, H.; Cloutier, D.; Shen, L.; Arizpe, A.; Gupta, S. V.; Fanget, M.; Seu, L.; Cathcart, A.L.; Lau, A.H.; Hwang, C.; Gane, E. Reduction in Hepatitis B Viral DNA and Hepatitis B Surface Antigen Following Administration of a Single Dose of VIR-3434, an Investigational Neutralizing Monoclonal Antibody: First Experience in a Population With Viremia. American Association for the Study of Liver Diseases (AASLD). Washington, DC, USA; November 4–8, 2022.

[17] Burm R, Van Houtte F, Verhoye L, Mesalam AA, Ciesek S, Roingeard P, et al. A human monoclonal antibody against HBsAg for the prevention and treatment of chronic HBV and HDV infection. JHEP Rep 2023;5:100646.

[18] Li D, He W, Liu X, Zheng S, Qi Y, Li H, et al. A potent human neutralizing antibody Fc-dependently reduces established HBV infections. eLife 2017;6.

[19] Jiang Y, Chen X, Ye X, Wen C, Xu T, Yu C, et al. A Dual-domain Engineered Antibody for Efficient HBV Suppression and Immune Responses Restoration. Adv Sci (Weinh) 2024;11:e2305316.

[20] Michler T, Zillinger J, Hagen P, Cheng F, Festag J, Kosinska A, et al. The lack of HBsAg secretion does neither facilitate induction of antiviral T cell responses nor Hepatitis B Virus clearance in mice. Antiviral Res 2024;226:105896.

[21] Ouaguia L, Leroy V, Dufeu-Duchesne T, Durantel D, Decaens T, Hubert M, et al. Circulating and Hepatic BDCA1+, BDCA2+, and BDCA3+ Dendritic Cells Are Differentially Subverted in Patients With Chronic HBV Infection. Front Immunol 2019;10:112.

[22] Gehring AJ, Haniffa M, Kennedy PT, Ho ZZ, Boni C, Shin A, et al. Mobilizing monocytes to cross-present circulating viral antigen in chronic infection. The Journal of clinical investigation 2013;123:3766–3776.

[23] Carotenuto P, Artsen A, Niesters HG, Osterhaus AD, Pontesilli O. In vitro use of autologous dendritic cells improves detection of T cell responses to hepatitis B virus (HBV) antigens. J Med Virol 2009;81:332–339.

[24] Ma YJ, He M, Han JA, Yang L, Ji XY. A clinical study of HBsAg-activated dendritic cells and cytokine-induced killer cells during the treatment for chronic hepatitis B. Scand J Immunol 2013;78:387–393.

[25] Michler T, Kosinska AD, Festag J, Bunse T, Su J, Ringelhan M, et al. Knockdown of Virus Antigen Expression Increases Therapeutic Vaccine Efficacy in High-Titer Hepatitis B Virus Carrier Mice. Gastroenterology 2020;158:1762–1775.e1769.

[26] Mousavi SA, Sporstol M, Fladeby C, Kjeken R, Barois N, Berg T. Receptor-mediated endocytosis of immune complexes in rat liver sinusoidal endothelial cells is mediated by FcgammaRIIb2. Hepatology (Baltimore, Md) 2007;46:871–884.

[27] Ganesan LP, Kim J, Wu Y, Mohanty S, Phillips GS, Birmingham DJ, et al. FcgammaRIIb on liver sinusoidal endothelium clears small immune complexes. J Immunol 2012;189:4981–4988.

[28] Schurich A, Bottcher JP, Burgdorf S, Penzler P, Hegenbarth S, Kern M, et al. Distinct kinetics and dynamics of cross-presentation in liver sinusoidal endothelial cells compared to dendritic cells. Hepatology (Baltimore, Md) 2009;50:909–919.

