## Supplemental methods and data for "The engineered monoclonal antibody tobevibart enhances HBsAg capture by Fc receptor positive cells leading to activation of HBV-specific T cells"

#### Supplemental Information Online

### Supplementary Material and Methods

##### Ethics statement for the use of human samples

Whole blood, PBMCs, sera or plasma samples were obtained from human subjects under study protocols approved by the local Institutional Review Boards (Ethics Committees of the Canton Ticino, Switzerland or IRB of the University of California San Francisco or Advarra IRB for Quest Diagnostics, USA). All donors provided written informed consent for the use of blood and blood components.

##### Ethics statement and clinical trial

VIR-3434-1002 is a randomized, double-blind, placebo-controlled, phase 1, single-ascending-dose study (ClinicalTrials.gov, NCT04423393) examining the safety, tolerability, and antiviral activity of tobevibart (VIR-3434). Eligible participants were adults (≥18 years of age) with chronic HBV infection and without cirrhosis. This clinical trial was designed, conducted and funded by Vir Biotechnology, Inc. as sponsor. The study was conducted in accordance with consensus ethical principles derived from international guidelines, including the Declaration of Helsinki and Council for International Organizations of Medical Sciences, applicable International Council for Harmonization and Good Clinical Practice and applicable laws and regulations. Approval from the local Institutional Review Board or Independent Ethics Committee was obtained and informed consent was obtained from all participants prior to their participation in the study. Here, we report data from an exploratory analysis using stabilized whole blood samples of participants receiving a single, subcutaneous dose of 300 mg tobevibart.

##### HBsAg production from PLC/PRF/5 cells

HBsAg was produced using PLC/PRF/5 cells (Alexander cells, hepatoma cell line, Sigma) cultured in a CELLine two compartment bioreactor flask (SIGMA, Z688045). Briefly, cells were resuspended at $1x10^6$ cells/ml in 15 ml complete medium (DMEM high glucose with glutamine, 10% FBS), and seeded in the cell compartment, which is separated from the medium compartment (containing 950 mL complete media) by a 10 kDa semi-permeable cellulose acetate membrane. Samples were incubated at 37°C for 21 days, at which point media from the cell compartment was harvested and centrifuged at 400g for 4 minutes prior to HBsAg quantification

##### HBsAg quantification

HBsAg produced by PLC/PRF/5 cells or serum samples from patients with CHB (source: BioIVT) were quantified using the diagnostic HBsAg quantitative electrochemiluminescence immunoassays (ECLIA): Elecsys HBsAg II quant II (Cobas e801, Roche, 07027443119) or Abbott Architect HBsAg quant (i2000SR, 6C36), with a LLOQ (lower limit of quantification) = 0.05 IU/mL. For quantification of endogenous HBsAg within whole blood from patients with CHB (from cohorts outside the VIR clinical studies and that were collected by the University of California San Francisco or Quest Diagnostics, USA), HBsAg levels were quantified using the Elecsys HBsAg II quant II assay (Cobas e801, Roche, 07027443119) on matched plasma samples collected at the same time as the whole blood samples used for the *in vitro* ICs binding assay. For Cytodelics stabilized whole blood samples collected from patients with CHB enrolled in a phase 1 clinical trial and receiving a single dose of 300 mg tobevibart, HBsAg levels of serum samples that matched time points of whole blood collection were quantified using the Abbott Architect assay.

#### Electron Microscopy

Preparation of Negative-Stained Specimens: HBsAgadw was obtained from Prospec (HBS-872). Two concentrations of HBsAg were incubated alone or with a specified concentration of tobevibart (VIR-3434) in a 37°C water bath for 60 minutes (see Fig. 1 for concentrations). HBsAg and tobevibart were kept at 4°C before mixing and immediately placing in water bath. Dilutions were made with PBS. A negatively stained specimen of each sample was prepared on ultrathin amorphous carbon supported on a 200-mesh Cu grid. (The carbon-coated grid was first placed in a lab-made plasma, glow-discharge device for 10-15 seconds to clean the carbon and render it more hydrophilic.) Stain solution was 1% ammonium molybdate. Specimens were prepared using the following steps: within 0.5–1 minute of removal from 37°C water bath, application of 3.5 µL sample (undiluted) to the carbon-coated grid for one minute then blot via filter paper; rapid 2x application of 20 µL PBS then blotting immediately after each application (4–5 sec total); rapid 2x application of stain solution then blotting immediately after each application (4–5 sec total); final application of 20 µL stain solution for 15–20 seconds, blot, dry in air, then vacuum for 21–43 minutes. Imaging of Negative-Stained Specimens: Grids were imaged on a ThermoFisher Tecnai 12 transmission electron microscope (TEM) operated at 120 kV. At least 30 images of each specimen were recorded at different positions across each grid using a Gatan UltraScan camera. A nominal magnification setting of 30,000× (calibrated magnification = 41,290[±60]×) was used, which corresponded to 3.39 (±0.01) Å/pixel. Images were recorded between 0.5 and 2.1 µm underfocus, with an average and standard deviation of 1.2 (±0.2) µm. Representative images were selected and the approximate ratio of tobevibart (VIR-3434) and HBsAg leading to a peak immune complex size was determined by evaluating all acquired pictures.

#### HBsAg binding ELISA

Half-area 96-well plates were coated with 25 µl/well HBsAg adw (Prospec, hbs-872-c) at 1 µg/mL and incubated overnight at 4°C. Plates were washed 3 times with PBS-T using an automated washer. Blocking solution (PBS with 1% BSA) was added in 100 µL/well and plates were further incubated for 90 min at room temperature (RT). Blocking solution was removed and plates were washed again 3 times. Then 25 µL/well of serial 1:3 mAb dilutions (1 µg/mL to 0.017 ng/mL) in blocking buffer were dispensed and plates were incubated 90 min at RT. Plates were then washed 4 times with PBS-T (220 µl/well). The HRP-conjugated secondary antibody reagent goat anti-human IgG (Jackson Immuno, 109-036-098) was added in 25 µL to each well at 0.16 µg/mL in blocking buffer and further incubated for 1 hour at RT. After 4 washes with PBS-T, 40 µl/well of Sureblue (TMB) ELISA substrate solution was dispensed in each well and plates were developed for 4-5 min at RT. The reaction was stopped with 40 µL/well of 1% HCl, and the OD was read at 450 nm in an ELISA reader (Bio-Tek, ELx808).

#### HBV neutralization (PHH)

Primary human hepatocytes (PHH, Thermo Fisher Scientific) were seeded at 58,000 cells/well in collagen-coated 96-well plates according to the manufacturer's instructions. Five hours post seeding, at ~90% confluence, infection was initiated. Infection mix was prepared by diluting the concentrated HBV stock virus at 1:30 in PHH maintenance medium (Williams E, primary hepatocyte maintenance supplements, Thermo Fisher Scientific, 2% FBS, 2% DMSO). Five-fold, eight-step serial dilutions of the antibodies were prepared in PBS starting at 62.5 µg/mL and diluted 1:10 into the infection mix. Infection control wells contained infection mix with PBS alone. Virus-antibody complexes were allowed to form by 30-minute incubation at 37°C, then 40% PEG-8000 was added to a final PEG concentration of 4% and mixed well. One hundred microliters from each well of the virus-antibody complex plates were pipetted in triplicates onto the cells. One day post infection, the inoculum was removed, cells were washed with PBS and fresh PHH maintenance medium was added. Medium was exchanged every 2-3 days post infection. At day 7 post infection, viral markers in the cell supernatant were quantified by HBeAg/HBsAg CLIA according to the manufacturer's instructions (Autobio Diagnostics).

#### mAb binding to human FcgRs via surface plasmon resonance (SPR)

Binding of HBC34* variants to human FcγRs was measured by SPR using a Biacore T200 instrument. Ligands and analytes were diluted in HBS-EP+ pH 7.4 (Cytiva) and all experiments were run at 25°C. Recombinant Avi-tag biotinylated huFcγRs (Acro) were injected over a CAP sensor chip as ligand. Increasing concentrations of HBC34* Fc variants were then injected over the functionalized sensor surface using single-cycle kinetics. Sensorgrams underwent double reference subtraction first from the reference flow cell and second from a buffer injection cycle. Steady-state affinities were determined by plotting equilibrium RU values (determined by averaging a 5 second window centered 5 seconds before the end of the association phase) versus analyte concentration for each injection. Rmax was fixed for all analytes for each receptor based on the Rmax value determined for the highest affinity binder in each set. For huFcyRIIb, a separate positive control antibody with the EFTAE (S267E, H268F, S324T, G236A, I332E) Fc mutations [1] was used to determine Rmax. Data were analyzed using Biacore Insight software version 4.0 (Cytiva) and visualized with GraphPad Prism (v10.1).

#### FcRn and C1q binding via Octet

Binding of HBC34-v35, HBC34-v35-LS or tobevibart (HBC34-v35-LS-GAALIE) to human FcRn (at pH 6.0 or 7.4) and binding of HBC34-v35-LS or tobevibart to C1q (Sigma) were measured on an Octet RED96 instrument (biolayer interferometry, BLI, ForteBio). Biosensors (Anti-Penta-HIS) pre-hydrated in kinetic buffer for 10 min at RT were loaded with human FcRn at 1 µg/mL for 8 min or with C1q at 3 µg/mL for 4 min in kinetics buffer. The baseline was measured in kinetics buffer at pH=6.0 or pH=7.4 for 1 minute for FcRn and in kinetics buffer at pH=7.1 for 1 minute for C1q. FcRn-loaded sensors were then exposed for 5 minutes to a solution of human mAb (HBC34-v35, HBC34-v35-LS, or tobevibart) 13.33 nM (equivalent to 2 µg/mL) in kinetics buffer at pH=7.4 or pH=6.0 to measure association of FcRn–mAb in different milieus (on rate). On the other hand, C1qloaded sensors were exposed for 10 minutes to a solution of human mAb (HBC34-v35-LS, or tobevibart) at 10 μg/mL in kinetics buffer at pH=7.1. Dissociation was then measured in kinetics buffer at the same pH for an additional 5 minutes (off rate) for FcRn and for an additional 4 minutes for C1q. All steps were performed while stirring at 1,000 rpm at 30°C. Association and dissociation profiles were measured in real time as change in the interference patterns.

##### Antibody-dependent signaling via human FcγRIIa and FcγRIIIa using Jurkat reporter cells

mAbs were serially diluted 4-fold in assay buffer, starting from 100 μg/mL to 0.00153 μg/mL for FcγRIIa, and 2.5-fold starting from 50 μg/mL to 0.0328 μg/mL for FcγRIIIa. Target antigen (HBsAg adw derived from Pichia pastoris, Prospec) was added in a white flat bottom 96-well plate in 25 μL at a final concentration of 1.14 μg/mL (corresponding to 250 IU/mL) for FcγRIIa, and at 22.72 μg/mL (corresponding to 5000 IU/mL) for FcγRIIIa. Then, serially diluted antibodies were added in 25 μL to each well, and the antigen/antibody mixture was incubated for 30 minutes at room temperature. Effector cells for the FcγRIIa or FcγRIIIa activation bioassay were thawed and added at a cell density of 5 x $10^4$/well and 7.5 x $10^4$/well, respectively, in 25 μL medium. Negative control wells contained either HBsAg and effector cells but no antibody or assay buffer only. Plates were incubated for 20 hours (FcγRIIa) or 24 hours (FcγRIIIa) at 37°C with 5% CO2. Activation of human FcγRIIa (H131 allele) or FcγRIIIA (F158 allele) in this bioassay results in the NFAT-mediated expression of the luciferase reporter gene. Luminescence was measured with a luminometer using the Bio-Glo-TM Luciferase Assay Reagent according to the manufacturer's instructions. The FcγRIIa activation data are expressed as the average of relative luminescence units (RLU) over the background by applying the following formula: (RLU at concentration x of mAbs – RLU of background).

##### Tobevibart-dependent signaling via human FcγRIIa and FcγRIIIa using Jurkat reporter cells in a matrix of mAb and HBsAg concentrations *in vitro*

HBsAg produced in PLC/PRF/5 Alexander cells or HBsAg from patient serum were serially diluted 3-fold in assay buffer from 1'000 to 4.12 IU/mL for FcγRIIa, and from 10'000 to 41.2 U/mL for FcγRIIIa. Simultaneously VIR-3434 was serially diluted 5-fold in assay buffer from 100 μg/mL to 0.000256 μg/mL for FcγRIIa, and 4-fold from 100 μg/mL to 0.00153 μg/mL for FcγRIIIa. Serial dilutions of the target antigens were added in a white flat bottom 96-well plate in 25 μL, then serially diluted tobevibart was added in 25 μL per well, and the antibody/antigen was incubated for 20 minutes at room temperature. Effector cells for the bioassays were thawed and added at a cell density of 5 x $10^4$/well or 7.5 x $10^4$/well in 25 μL. Plates were incubated for 26 hours or 18 hours respectively at 37°C with 5% CO2. Luminescence in these reporter cell assays were measured as described above.

##### Flow cytometric analysis of binding of HBsAg-mAb ICs to primary human immune cells in whole blood samples from patients with CHB *in vitro*

Fresh whole blood (6-8 mL) was collected from human donors with CHB (n=21, by the University of California San Francisco or Quest Diagnostics, USA from cohorts outside the VIR clinical studies) (n=11 female, n=10 male) into vacutainers containing sodium citrate (CPT) or acid citrate dextrose (ACD). Samples with matched serum quantification with HBsAg <LLOQ were excluded from the analysis. HBC34*-GAALIE or other Fc variants (GRLR, WT, GA or afuc) were added to fresh whole blood samples within 5 hours of collection. Zero or 50 µg/mL HBC34*-GAALIE or other Fc variants were mixed with 200 µL of whole blood from donors with CHB and samples were incubated at 37°C, 5% $CO_2$ for 2 hours. After incubation, whole blood was gently mixed with 200 µL of room-temperature Cytodelics Whole Blood Cell Stabilizer and incubated at room temperature for 10 minutes. Samples were then stored at -80°C until processing. Cytodelics-stabilized whole blood was processed according to manufacturer's instructions. Briefly, frozen Cytodelics-stabilized whole blood samples were thawed, fixed with Cytodelics Fixation Buffer for 15 minutes at room temperature, and then red blood cells were lysed with Cytodelics Lysis Buffer for 5-10 minutes at room temperature. Lysed samples were centrifuged and washed with Cytodelics Wash Buffer. The remaining leukocytes were then stained to prepare for flow cytometry **(Table S1).** First, FcγRs of cells were blocked to prevent unspecific antibody and/or IC binding using TruStain FcX for 20 minutes at room temperature and washed with FACS Buffer. To identify immune cell subsets, treated cells were stained in Brilliant Stain Buffer with the antibodies against cell surface markers in **Table S1** for 30 minutes at room temperature. To detect HBsAg in ICs on the cell surface also in the presence of HBC34* ICs, an anti-HBs mAb (HBi), which binds to the antigenic loop of HBsAg but does not compete with HBC34* binding to HBsAg, was used. MPE8 is an irrelevant antibody that was used to control for non-specific binding of HBi (both isotype rIgG1). Cells were washed twice with FACS Buffer (PBS + 2% FBS) and then acquired on the BD FACSymphony A5. To determine FcγR expression across different immune cells, another set of untreated cells (without mAb spiked-in) were stained in Brilliant Stain Buffer for 30 minutes at room temperature. Cells were washed twice with FACS Buffer (PBS + 2% FBS) and then acquired on the BD FACSymphony A5.

**Table S1**

| Reagent | Source | Catalog # |
|---|---|---|
| TruStain FcX | BioLegend | 422302 |
| MPE8-rIgG1 PE (control antibody that binds to an irrelevant antigen) | Vir Biotechnology, Bellinzona, Switzerland | N/A, Lot: 20180327/FAD |
| HBi-rIgG1 AF647 (anti-HBs mAb) | Vir Biotechnology, Bellinzona, Switzerland | N/A, Lot: 20121212/9XY |
| CD16 PE-Cy7 (clone 3G8) | BD Biosciences | 557744 |
| CD14 BUV805 | BD Biosciences | 612902 |
| CD19 BUV563 | BD Biosciences | 741361 |
| CD3 PE-CF594 | BD Biosciences | 562280 |
| CD56 BUV395 | BD Biosciences | 563554 |
| CD123 BUV737 | BD Biosciences | 741769 |

| Reagent | Source | Catalog # |
|---|---|---|
| CD11c BV711 | BD Biosciences | 563130 |
| HLA-DR APC-H7 | BD Biosciences | 561358 |
| CD32a FITC (clone IV.3) | StemCell | 60012FI |
| CD32b (2B6-muIg2a) AF647 (clone 2B6) | Vir Biotechnology, Bellinzona, Switzerland | N/A, Lot 20140502/PCX |
| HLA-DR PE-CF594 | BD Biosciences | 562304 |
| CD3 PE-Cy5.5 | eBioscience | 35-0036-42 |

Antibody Conjugation: The anti-CD32b antibody was conjugated to Alexa Fluor 647 per manufacturer's instructions (ThermoFisher). Briefly, 76 µg of anti-CD32b was conjugated to 1 vial of AF647 for 1 hour at room temperature and then purified using the purification resin. MPE8 was conjugated to PE and HBi conjugated to AF647 per manufacturer's instructions (Abcam). 100 µg of MPE8 or HBi was mixed with PE or AF647, respectively, incubated for 2.5 hours at room temperature, then quenched for 10 min. Conjugated HBi and MPE8 were buffer exchanged into PBS using a 50 kDa cutoff Amicon Centrifugal Filter Unit.

For both HBsAg IC detection and FcγR expression analysis, the following immune populations were identified: neutrophils (SSC-$A^{hi}$CD$16^{hi}$ or CD15+CD$16^{hi}$), B cells (non-neutrophils, HLA-DR+CD3-CD19+), classical monocytes (non-neutrophils, HLA-DR+CD3-CD19-CD14+CD16-), CD16+ non-classical (NC) monocytes (non-neutrophils, HLA-DR+CD3-CD19-CD14-CD16+), classical dendritic cells (cDCs, non-neutrophils, HLA-DR+CD3-CD19-CD14-CD16-CD123-CD11c+), plasmacytoid dendritic cells (non-neutrophils, HLA-DR+CD3-CD19-CD14-CD16-CD123+CD11c-), basophils (non-neutrophils, HLAD-CD3-CD56-CD123+), and CD16+ NK cells (non-neutrophils, HLADR-CD3-CD123-CD56+CD16+).

**Phase 1 clinical study of a single dose of 300 mg tobevibart in patients with CHB and detection of HBsAg ICs on circulating immune cells**

VIR-3434-1002 was a phase 1, randomized, double-blind, placebo-controlled, single ascending dose clinical study examining the safety, tolerabiltiy, and antiviral activity of tobevibart [2]. In Part B, a single dose of either 6 mg, 18 mg, 75 mg, or 300 mg tobevibart was administered in virally suppressed participants who are HBVe antigen (HBeAg) negative with HBsAg < 3,000 IU/mL. In Part C, a single dose of either 18 mg, 75 mg, or 300 mg tobevibart was administered in virally suppressed participants who are HBVe antigen (HBeAg) negative with HBsAg ≥ 3,000 IU/mL (except the 18-mg cohort, wherein no baseline HBsAg was specified). In Part D, a single dose of either 75 mg or 300 mg tobevibart was administered in participants who are viremic (HBV DNA ≥ 1,000 IU/mL) with any HBsAg level. In each cohort, participants were randomly assigned 6:2 to receive either tobevibart or placebo subcutaneously.

Tobevibart-mediated binding of endogenous HBsAg in ICs to immune cells was analyzed in a pre-dose (Day 1) sample and post-dose days 8, 15, 29, and 57. Analysis criteria was ≥500 events collected, and day 1 samples from each participant must be analyzable, which was the case for n = 17 out of total 18 participants dosed with a single dose of 300 mg tobevibart.

Whole blood samples were collected and added to Cytodelics Stabilizer in a 1:1 ratio in pre-filled tubes. After 10 minutes incubation at room temperature, samples were transferred to -70°C for storage. Samples were analyzed at Precision for Medicine. For processing, samples were thawed, cells were centrifuged, the supernatant aspirated, and cells incubated with Human TruStain FcX block for 30 minutes at RT to block non-specific binding. After the blocking step, the cells were washed twice with staining buffer. All aspiration steps were performed manually, with no decanting throughout the entire procedure to prevent the loss of cells. After the cells were washed in staining buffer, they were stained with an antibody mix (**Table S2**) against cell surface markers for 30 minutes at RT in the dark. Fluorescence minus-X (FMX) controls were stained only for the markers listed below (**Table S2**). After the surface staining, the cells were washed twice with staining buffer and then resuspended in 300 µL staining buffer for sample acquisition. The samples were acquired on a BD FACSymphony A5 flow cytometer, using predetermined BD FACS Diva software (version 9.0) application settings and MFI tracking with CS&T beads. Data analysis was performed using FlowJo software (version 10). Immune cell subsets were identified using the lineage markers and gating strategy listed in **Table S3**. For the detection of HBsAg ICs on the surface of immune cells, samples were stained with HBi, as above, an anti-HBs mAb that does not compete with tobevibart binding to HBsAg. HBi gate placement for each participant was determined using day 1 (pre-dose) samples and applied to post-dose samples. “ICs detected” was classification as ≥5% HBD7+ cells for any immune cell subset.

**Table S2: Antibody staining mix for cell surface markers (phase 1 clinical sample analysis)**

| Manufacturer | Catalog # | Fluorophores | Clone | Antibody | Function | Stain | FMX |
|---|---|---|---|---|---|---|---|
| Conjugation | N/A | AlexaFluor647 | HBD7 | HBi | Detect IC | √ | x |
| BD | 566482 | BB700 | 7G3 | CD123 | Lineage | √ | √ |
| BD | 740287 | BUV395 | HIB19 | CD19 | Lineage | √ | √ |
| BD | 741187 | BUV496 | W6D3 | CD15 | Lineage | √ | √ |
| BD | 741394 | BUV563 | IA6-2 | IgD | Subset | √ | √ |
| BD | 741837 | BUV737 | HIT2 | CD38 | Subset | √ | √ |
| BD | 612902 | BUV805 | M5E2 | CD14 | Lineage | √ | √ |
| Biolegend | 329920 | BV421 | EH12.2H7 | PD1 | Inhibitory | √ | x |
| BD | 563092 | BV510 | L128 | CD27 | Subset | √ | √ |
| Biolegend | 302040 | BV605 | 3G8 | CD16 | Lineage | √ | √ |
| BD | 563412 | BV650 | FUN-1 | CD86 | Activation | √ | x |
| BD | 563130 | BV711 | B-Ly6 | CD11c | Lineage | √ | √ |

| | | | | | | | |
|---|---|---|---|---|---|---|---|
| BD | 747068 | BV750 | NCAM16.2 | CD56 | Lineage | √ | √ |
| BD | 740984 | BV786 | GA-R2 | CD235a | Dump | √ | √ |
| Biolegend | 307604 | FITC | L243 | HLADR | Lineage | √ | √ |
| Biolegend | 305322 | PE | HB15e | CD83 | Activation | √ | x |
| BD | 551128 | PE-Cy5 | 1C6 | CXCR3 | Homing | √ | x |
| Invitrogen | 3500364 | PE-Cy5.5 | SK7 | CD3 | Lineage | √ | √ |
| Biolegend | 314532 | PE-Cy7 | MHM-88 | IgM | Subset | √ | √ |
| Biolegend | 354922 | PE-Dazzle594 | Bu32 | CD21 | Subset | √ | √ |

**Table S3: Gating for identification of immune cell subsets (phase 1 clinical sample analysis)**

| Cell population | Source |
|---|---|
| Neutrophils | CD15+ CD16+ |
| CD16+ NK cells | CD15– CD16– CD3– CD123– CD56+ CD16+ |
| CD16+ monocytes | CD15– CD16– HLA-DR+ CD56– CD19– CD14+/– CD16+ |
| Classical monocytes | CD15– CD16– HLA-DR+ CD56– CD19– CD14+ CD16– |
| B cells | CD15– CD16– HLA-DR+ CD56– CD19+ |
| cDCs | CD15– CD16– HLA-DR+ CD56– CD19– CD14– CD16– CD11c+ CD123– |

##### Competition ELISA to determine non-competition between HBC34* and HBi in binding to HBsAg

Half-area 96-well plates were coated with 25 µL/well HBsAg adw at 1 µg/mL and incubated overnight at 4°C. Then 25 µl/well of serial 1:4 competitor non-biotinylated mAb dilution of HBC34 or HBi (25 µg/mL to 0.024 µg/mL) were dispensed. Then without washing the plates, 25 µl/well of serial 1:4 dilution of the other biotinylated mAb HBi or HBC34, respectively, (25 µg/mL to 0.024 µg/mL) were dispensed on top. Plates were then incubated for 1.5 hour at RT. The AP-conjugated streptavidin was added in 50 µL to each well and further incubated for 45 minutes at RT. After addition of substrate solution OD was read at 405 nm in an ELISA reader. For data analysis OD were converted to % inhibition.

##### Primary human monocyte isolation from buffy coats and differentiation to moDCs

Human mononuclear cells (PBMCs) were isolated from peripheral blood buffy coats of healthy donors (obtained from the Swiss Blood Donation Center of Lugano, Switzerland) using Ficoll-Paque PLUS and density gradient centrifugation. Monocytes (CD14+ cells) were further isolated by positive selection using magnetic microbeads. Monocytes were cultured in complete medium (RPMI 1640, 10% FBS, 1% NEAA, 1% Glutamine, 1% Pen/Strep, 1% Sodium Pyruvate, β-mercaptoethanol 50µM) in a 12-well plate flat bottom at a concentration of 500,000 cells/mL. To induce the differentiation of monocytes into monocyte-derived dendritic cells (moDCs), the complete medium was enriched by a cocktail of cytokines including GM-CSF (50 ng/mL) and IL-4 (1,000 IU/mL). Monocytes were differentiated into immature moDCs for 6 days.

##### Stimulation of human moDCs with HBsAg-mAb ICs and assessing their activation by Flow Cytometry

Each stimulating condition was prepared in a 96-well round bottom plate, containing HBC34*-GAALIE or other Fc variants (WT, GA or GRLR, 50 µg/mL) in combination with HBsAg (30-1000 IU/mL). The plate was then incubated at 37 °C for at least 1 hour to allow the formation of ICs between HBsAg and mAbs. As controls, wells contained only mAb (50 µg/mL), only HBsAg (30-1000 IU/mL) or medium alone. As source of HBsAg, the serum of patients with CHB was used **(Table S4)**. LPS (100 ng/mL) was used as positive control of moDCs activation. 100,000 or 200,000 moDCs were gently added to each well. moDCs together with immune complexes were incubated at 37 °C for 21-24 hours before assessing moDCs activation. Cells were stained with antibodies against CD14, CD83, CD86 and HLA-DR conjugated to fluorochromes **(Table S5).** To determine cell viability cells were stained with Zombie Aqua. Data were acquired using the ZE5 Cell Analyzer (Bio-Rad).

**Table S4: List of serum samples from patients with CHB**

| **Sample set** | **Lot** | **Source** |
|---|---|---|
| HMN421933 | 118486 | BioIVT |
| HMN432911 | 119487 | BioIVT |
| HMN555659 | 213-0476 | BioIVT |
| HMN555661 | 213-0481 | BioIVT |
| HMN555666 | 47-2308 | BioIVT |
| HMN69009 | 101131 | BioIVT |
| HBsAg from PLC cells | 16-Sep-2019 | In house production |

HBsAg was quantified using the diagnostic HBsAg quantitative assays Elecsys HBsAg II quant II (Cobas e801, Roche, Cat# 07027443119) or Abbott Architect HBsAg quant (i2000SR, Cat# 6C36), with a LLOQ = 0.05 IU/mL.

**Table S5: List of used anti-human mAbs for surface staining**

| **Item** | **Source** | **Catalog #** |
|---|---|---|
| CCR7 (PE) | Lucerna Chem | 353204 |
| CD14 (APC) | BD Biosciences | 561708 |
| CD25 (PerCP-Cy5.5) | Lucerna Chem | 302626 |
| CD3 (BV711) | Lucerna Chem | 317328 |

| | | |
|---|---|---|
| CD4 (APC-Cy7) | BD Biosciences | 557871 |
| CD8 (BV605) | Lucerna Chem | 300936 |
| CD45RA (AF488) | Lucerna Chem | 304114 |
| CD45RA (PE-Cy7) | Lucerna Chem | 304126 |
| CD83 (BV421) | Lucerna Chem | 305324 |
| CD86 (PE-Cy7) | Lucerna Chem | 305422 |
| HLA-DR (FITC) | Lucerna Chem | 307603 |
| HLA-DR (Pacific Blue) | Lucerna Chem | 307633 |
| HLA-DR (APC) | Lucerna Chem | 307610 |
| ICOS (BV421) | BD Biosciences | 562901 |

**Quantification of cytokines in human moDCs culture supernatants**

After 21-24 hours of moDCs stimulation by HBsAg ICs, the supernatant was harvested and a set of 10 cytokines (IFN$\gamma$, IL-1b, IL-2, IL-4, IL-6, IL-8, IL-10, IL-12p70, IL-13, and TNFα) was quantified using the V-PLEX Proinflammatory Panel 1 Human Kit (MSD) following the manufacturer's procedure. Electro chemiluminescence (ECL) signal was recorded using the cooled CCD camera of the QuickPlex SQ 120 Instrument (MSD), and Discovery Workbench 4.0 (MSD) was used to extrapolate cytokine concentrations from ECL signals.

**Stimulation of moDCs from human HBV vaccinees and co-culture with autologous CD4+ memory T cells**

PBMCs were freshly isolated from whole blood of healthy human HBV vaccinees following the protocol described above. Monocytes (CD14+ cells) were further isolated by positive selection using magnetic microbeads and differentiated into moDCs for 6 days with GM-CSF and IL-4 as described above. For a detailed listing of used reagents, see **Table S6**. The negative fraction (PBMCs minus monocytes) was stored in liquid nitrogen for the subsequent isolation of CD4+ memory T cells.

Each stimulating condition was prepared in a 96-well flat bottom plate, containing HBC34*-GAALIE or other Fc variants (WT, GA or GRLR, 50 µg/mL) in combination with HBsAg (30 or 100 IU/mL). The plate was then incubated at 37 °C for at least 1 hour to allow the formation of immune complexes between HBsAg and mAbs. Negative controls contained media alone or HBsAg alone. 10,000 moDCs were gently added to each well and samples were incubated at 37 °C for 21-24 hours to allow for stimulation of moDCs prior to addition of CD4+ memory T cells.

The autologous frozen negative fraction (PBMCs minus monocytes) was thawed, total CD4+ T cells were enriched by positive selection using magnetic microbeads and then memory CD4+ T cells were sorted based on the expression of the surface markers: CD4+CD25–CD45RA– (CCR7+/–), (**Table S5**). CD4+ memory T cells were sorted using the Sony cell sorter SH800SFP and labelled with carboxyfluorescein succinimidyl ester (CFSE). Finally, 100,000 CFSE-labelled CD4+ memory T cells were put in co-culture with stimulated moDCs. The cells were incubated for 5 days at 37 °C. SEB (1 µg/mL) was used as positive control of T cell proliferation. To assess the expression of activation markers, CD4+ memory T cells were stained with anti-CD4, anti-CD25, anti-CD45RA, anti-CCR7, anti-ICOS, anti-HLA-DR antibodies conjugated to fluorochromes **(Table S5)**. To determine cell viability, cells were stained with Zombie Aqua. Data were acquired using the ZE5 Cell Analyzer (Bio-Rad).

**Table S6, List of used reagents for moDC and T cell cultures**

| **Item** | **Source** | **Catalog #** |
|---|---|---|
| CD14 MicroBeads, human | Miltenyi Biotec | 130-050-201 |
| CD4 MicroBeads, human | Miltenyi Biotec | 130-045-101 |
| Naive CD4+ T Cell Isolation Kit II, human | Miltenyi Biotec | 130-094-131 |
| CellTrace CFSE Cell Proliferation Kit | Thermo Fisher Scientific | C34554 |
| Lipopolysaccharides (LPS) from *Salmonella enterica* serotype typhimurium | Sigma-Aldrich Chemie | L2262-5MG |
| Recombinant Human GM-CSF | Miltenyi Biotec | 130-093-865 |
| Recombinant Human IL-4 | Bio-Techne | 204-IL-020 |
| Staphylococcal enterotoxin B (SEB) from *Staphylococcus aureus* | Sigma-Aldrich Chemie | S4881-1MG |
| Zombie Aqua Fixable Viability Kit | Lucerna Chem | 423101 |
| V-PLEX Proinflammatory Panel 1 Human Kit for multiplexed quantification of 10 cytokines | MSD | K15049D-2 |

##### Quantification of human FcγR expression on primary monocytes and moDCs

Human mononuclear cells (PBMCs) were isolated from peripheral blood buffy coats of healthy donors (obtained from StemCell or Charles River) and monocytes and moDC were isolated as described above. To allow direct comparison between the density of various FcγR expressed by CD14+ monocytes and moDC, we used Quantum Simply Cellular kit (Bangs Laboratories), as per manufacturer’s recommendation. To determine optimal concentration, titration of the following antibodies was performed: anti-CD32a (clone IV.3, FITC conjugated from StemCell), anti-CD32b (clone 2B6, generated in house and conjugated to AlexaFluor 647), anti-CD16 (clone 3G8, PE labeled from Biolegend) and anti-CD64 (clone 10.1, PE labelled from BD Biosciences). Subsequently, staining of cell samples (in the presence of Truestain Fc receptor blocker from Biolegend), and beads was performed with the various FcγR antibodies at saturation and analyzed by flow cytometry.

##### Transgenic T cells activation assays

To assess antigen specific T cell responses to immune complexes, a triple reporter Jurkat line (Rosskopf et al. Oncotarget 2018) was engineered to overexpress CD4 and a human TCR specific for HBsAg. Antigen specific stimulation was performed using HLA matched moDCs (from healthy donors) treated overnight with ICs of HBsAg and mAb Fc variants. ICs were generated by mixing mAbs with 1000 IU/mL of HBsAg, obtained from PLC/PRF/5 cells, for 30 minutes at room temperature prior to addition to moDCs. In some experiments blocking antibodies for anti-FcgRs (anti-CD16 clone B73.1, Invitrogen; anti-CD32a, clone IV.3, Bio X Cell InvivoMab; and anti-CD64, clone 10.1, BD Biosciences) were used. MoDCs were pretreated with 1 μg/mL anti-Fc gamma receptor antibodies for 1 h at 37C, prior to stimulation with ICs. Following moDCs stimulation, Jurkat HBsAg TCR cell lines were added at a 1:1 ratio overnight, and NFAT-GFP, NFκβ-CFP and AP1-mCherry reporter expression and CD69 upregulation were analyzed by flow cytometry.

##### Mouse immunization with Engerix B vaccine

Mice were bred and experiments were performed at the Institute of Research in Biomedicine, Bellinzona, Switzerland facility, strictly following local guidelines for animal welfare. The *in vivo* work was authorized by the local IACUC authority (license n. TI-42/2020). Eight- to 10-week-old WT C57BL/6 mice were primed with 2 μg HBsAg per mouse of the Engerix B20 vaccine (100 μL subcutaneously + 100 μL intraperitoneally). Two weeks post first immunization, HuFcγR mice were boosted with additional 2 μg per mouse of the Engerix B20 vaccine as above.

##### Mouse bone marrow (BM) isolation, differentiation into BMDCs, and stimulation with ICs containing HBsAg and HBC34*-GAALIE or Fc variants

Femurs of HuFcγR mice transgenic for the full set of human FcγRs (I, IIa-R, IIIa-F, and IIb) [3] were collected, and bone marrow (BM) was isolated by flushing 30 μL of PBS into the lumen of the bone. BM cells were centrifugated 5 minutes at 500g (4°C) and re-suspended in 1 mL of pre-warmed BMDCs differentiation medium. Seven million BM cells were then seeded in one 10-cm Petri culture dish after re-suspension with 10 mL of pre-warmed BMDCs differentiation medium and further incubated for 8 days at 37°C. After 8 days of incubation at 37°C, BMDCs were stained to assess the expression of activation marker CD86 and the increase in the expression of CD11c and MHC class II (I-Ab), which is characteristic of differentiation of monocytes into BMDCs. Briefly, cells were stained with Zombie Aqua for 30 min at RT to determine cell viability. After a washing step with FACS buffer and a centrifugation at 400g for 5 min (4°C), BMDCs were stained with a surface markers mix containing antibodies recognizing the markers mentioned above, conjugated to different fluorochromes (CD86-APC, CD11c-PE, I-Ab-biotinylated), and incubated for 20 minutes on ice. After a washing step with FACS buffer and a centrifugation at 400g for 5 min (4°C), cells were stained with Streptavidin-PE/Cy7 on ice for 10 minutes. Finally, the cells were washed with FACS buffer and centrifugated at 400g for 5 minutes at 4°C, re-suspended in FACS buffer, and acquired using the ZE5 Cell Analyzer (Bio-Rad). The experimental conditions for the stimulation *in vitro* of BMDCs from HuFcγR mice by HBsAg in ICs with HBC34* Fc variants (GAALIE, GRLR, WT, GA) were prepared in a volume of 100 μL complete medium in a 96-well, flat-bottom plate.

The used stimulation conditions were: 1) mAb alone (HBC34*-GAALIE or Fc variants (WT, GA or GRLR, 20 µg/mL), 2) HBsAg alone (1000 IU/mL), 3) ICs of HBC34*-GAALIE or Fc variants (WT, GA or GRLR, 20 µg/mL) and HBsAg (1000 IU/mL), complexed in the culture plate for 1 hour at 37°C. SEB (1 µg/mL) was used as positive control for T cell proliferation. LPS (100 ng/mL) was used as positive control for BMDCs activation. Fifty thousand BMDCs resuspended in 100 µL complete medium, were gently added to each well containing 100 µL complete medium with IC or control stimuli and incubated at 37 °C for 21-24 hours.

**Isolation of CD4+ memory T cells from immunized mice**

Two months post immunization, mice were euthanized to allow organ collection (axillary, brachial and inguinal lymph nodes and spleens). A single cell suspension was obtained via gently smashing the lymphoid organs with a 5-mL syringe plunger on 70 µm nylon cell strainer in 20 mL FACS buffer. The obtained cell suspension was pelleted at 400g for 5 min 4°C, resuspended with 5 mL ACK buffer for 5 min to eliminate red blood cells, washed once with FACS buffer, and finally re-suspended in complete medium. CD4+ T cells were isolated by negative selection using magnetic microbeads from the CD4+ T Cell Isolation Kit (MACS, Miltenyi). Briefly, 100 µL of biotin-antibody cocktail were added to the cell suspension and incubated 5 min on ice. Two hundred µL of anti-biotin microbeads were added and incubated 10 min on ice. After washing the cells with MACS buffer at 400g for 5 min at 4°C, the pellet was resuspended in 3 mL of MACS buffer and added to the LS magnetic column attached to the QuadroMACS separator (Miltenyi). The LS column was detached from the magnet and the CD4+ T cells were eluted using MACS buffer. CD4+ CD44+ memory T cells then were sorted based on the expression of surface markers. Briefly, MACS-isolated CD4+ T cells were washed with FACS buffer and then resuspended in 200 µL of surface staining mix containing antibodies recognizing the markers mentioned above conjugated to different fluorochromes (CD4-APC-Cy7, CD44-PerCP) and incubated for 20 min on ice. After a washing step with FACS buffer and a spin at 400g for 5 min at 4°C, T cells were resuspended in FACS buffer, filtered with Pre-Separation Filters (30 µm) and sorted using the Sony cell sorter SH800SFP. To follow the proliferation of CD4+ memory T cells in co-culture with ICs-pulsed BMDCs, T cells were labeled for 8 min at 37°C with CFSE, at a final concentration of 5 µM in 1 mL of PBS, supplemented with 2% FBS. CD4+ T cells were then washed three times with complete medium at 300g for 8 min.

**Co-culture of ICs-pulsed BMDCs from HuFcγR mice with CD4+ Memory T cells from HBV-immunized mice**

CFSE-labeled CD4+ memory T cells from HBV-immunized mice were put in co-culture with BMDCs that were stimulated the day before, as described above. ICs-pulsed BMDCs in 96-well plate flat bottom were centrifuged at 450g for 5 min and the supernatant was discarded. BMDCs were then re-suspended in 200 µL of complete medium containing CFSE-labeled CD4+ memory T cells for a final co-culture condition of 1:10 (5E04 BMDCs and 5E05 CD4+ memory T cells). The co-coculture was incubated for 6 days at 37 °C. After 6 days, cells were stained to assess the CD4+ memory T cells activation and proliferation. Briefly, cells were stained with ZombieAqua for 30 min at RT to determine cell viability. After a washing step with FACS buffer and a spin at 400g for 5 min at 4°C, cells were stained with a mix containing antibodies against cell surface markers CD4-APC/Cy7, CD44-PerCP, CD69-Pe-Dazzle 594, CD62L-BV605 and were incubated for 20 min on ice. After a washing step with FACS buffer and a spin at 400g for 5 min at 4°C, cells were re-suspended in FACS buffer and acquired using the ZE5 Cell Analyzer (Bio-Rad). CD44+ CFSE low cells were interpreted as memory CD4+ T cells that were activated and had proliferated.

##### Data and statistical analyses

Flow cytometry data analysis was performed using Flowjo software. Other data were plotted and statistically analyzed using the GraphPad Prism 10.0 software.

#### Supplementary References

[1] Moore GL, Chen H, Karki S, Lazar GA. Engineered Fc variant antibodies with enhanced ability to recruit complement and mediate effector functions. MAbs 2010;2:181-189.

[2] Wong RG, Sneha V; Wang, Li; Camus, Gregory; Chen, Yi-Pei; Cathcart, Andrea L; Imam, Hasan; di Iulio, Julia; Momin, Amin; Aripze, Andre; Sun, David; Cloutier, Daniel; Arvin, Ann; Agarwal, Kosh; Yuen, Man-Fung; Wedemeyer, Heiner; Gane, Ed. Initial Evaluation of Immune Complex Binding in a Phase 1 Safety and Tolerability Study of Chronic HBV Participants Given a Single Dose of VIR-3434. AASLD; 2023.

[3] Smith P, DiLillo DJ, Bournazos S, Li F, Ravetch JV. Mouse model recapitulating human Fcgamma receptor structural and functional diversity. Proceedings of the National Academy of Sciences of the United States of America 2012;109:6181-6186.

#### Supplementary Figures and Legends

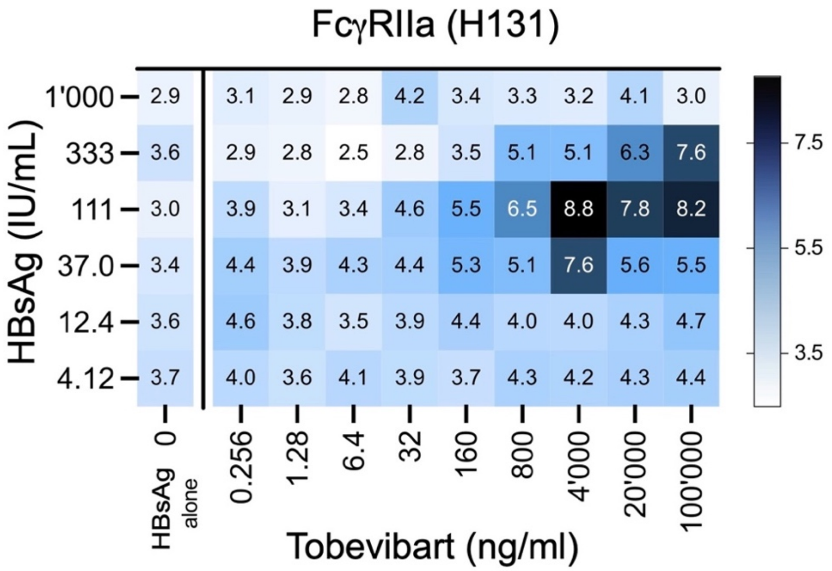

**Supplementary Figure S1: Tobevibart (VIR-3434) induces FcγRIIa signaling in ICs with HBsAg derived from HBV+ patient serum.**
Matrix of tobevibart and HBsAg concentrations assessed for FcγRIIa H131 signaling using Promega reporter cells. HBsAg was derived from serum of a patient with CHB.

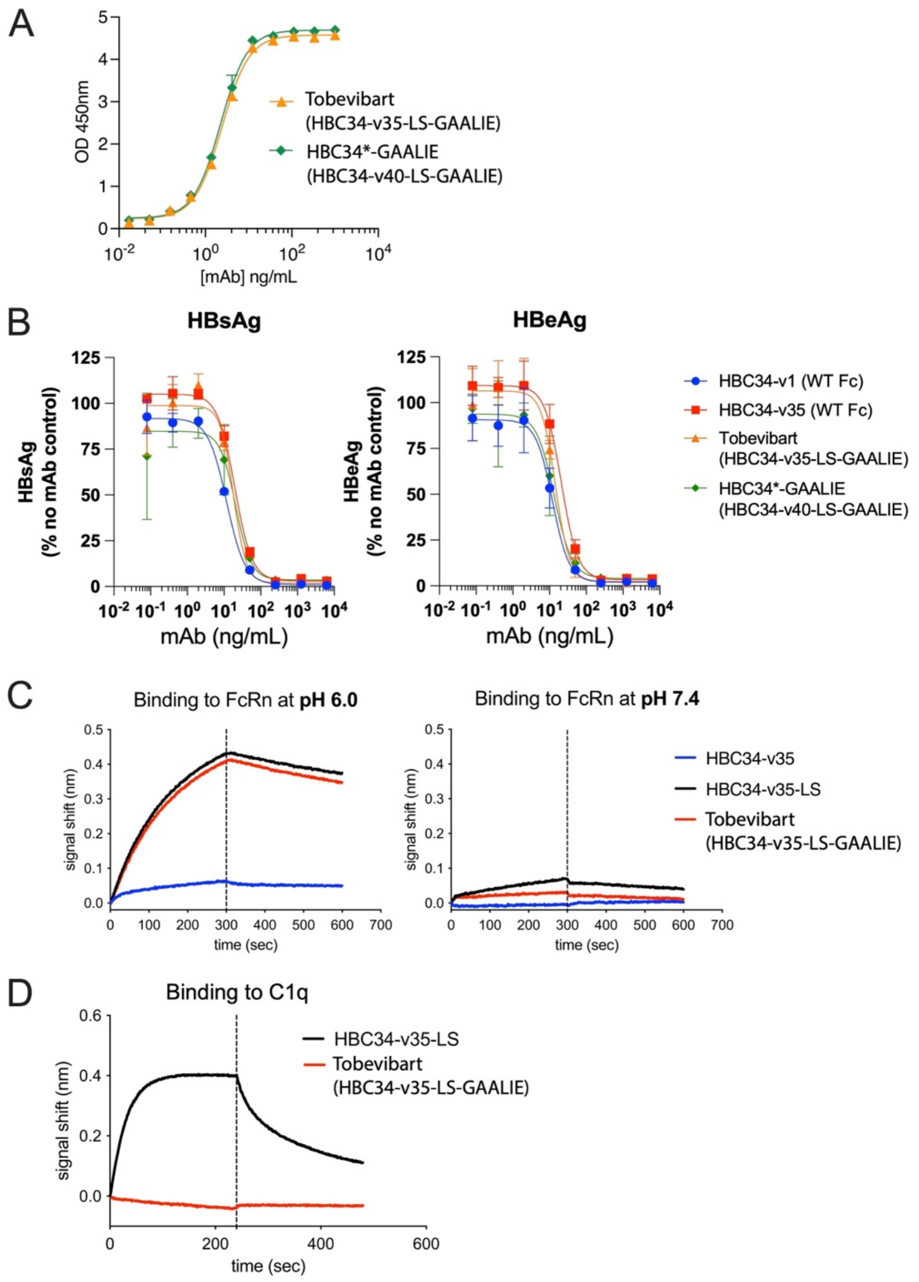

**Supplementary Figure S2: The Fab variants Tobevibart and HBC34*-GAALIE similarly bind HBsAg, neutralize HBV infection, and bind FcRn or C1q.**
**(A)** ELISA binding of Fab variants Tobevibart (HBC34-v35-LS-GAALIE) and HBC34*-GAALIE (HBC34-v40-LS-GAALIE) to HBsAg serotype adw.
**(B)** HBV neutralization of infection of primary human hepatocytes of Fab variants (HBC34-v1, -v35, -v40) and Fc variants (WT vs LS-GAALIE), measured via intracellular staining for HBsAg (left) or HBeAg (right).
**(C)** Binding of HBC34-v35 Fc variants (WT vs LS) or tobevibart (LS-GAALIE) in solution to immobilized human FcRn, measured via Octet biolayer interferometry in real time at pH=6.0 (left) or pH=7.4 (right). The time point 0 seconds represents switch from base line buffer to buffer containing human antibodies. Time point 300 seconds (dotted vertical line) represents switch to blank buffer at the corresponding pH. Curves indicate association and dissociation profiles of change in the interference patterns.
**(D)** Binding of immobilized HBC34-v35 Fc variants (LS) or tobevibart (LS-GAALIE) to C1q in solution via Octet.

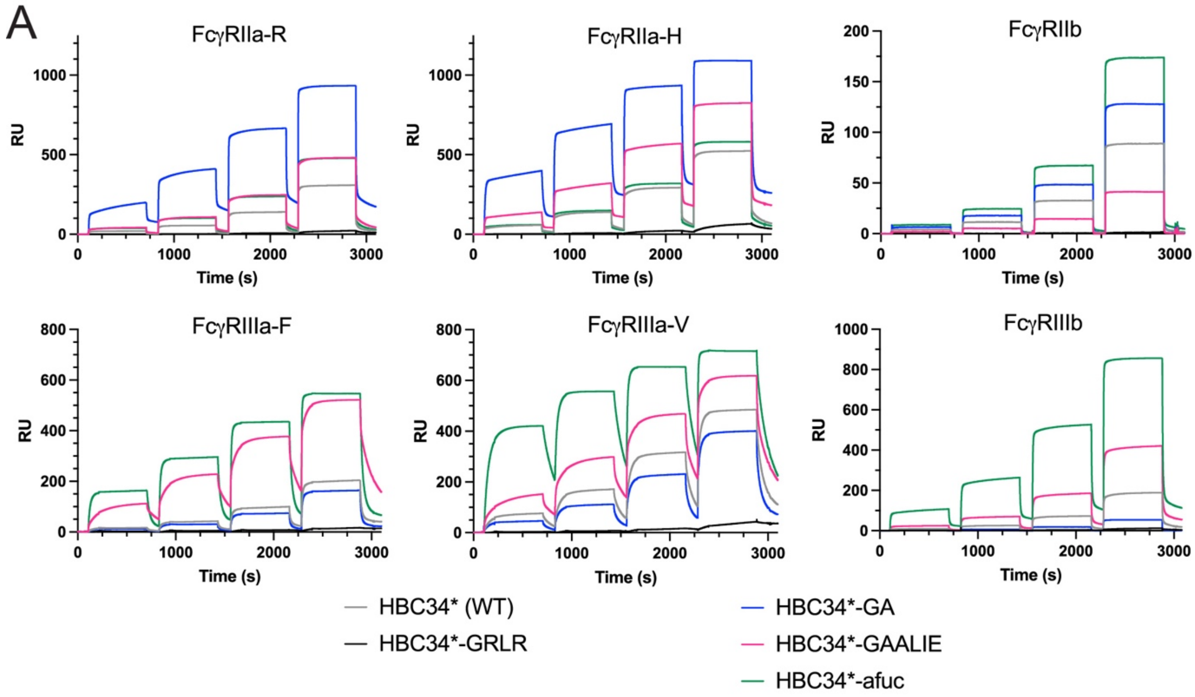

B

| mAb | Fold change to HBC34* (WT) FcγRIIa-R | FcγRIIa-H | FcγRIIb | FcγRIIIa-F | FcγRIIIa-V | FcγRIIIb |
|---|---|---|---|---|---|---|
| HBC34* (WT) | 1.00 | 1.00 | 1.00 | 1.00 | 1.00 | 1.00 |
| HBC34*-GA | 10.47 | 14.90 | 1.49 | 0.71 | 0.57 | 0.25 |
| HBC34*-afuc | 1.91 | 1.19 | 2.13 | 16.95 | 15.08 | 12.87 |
| HBC34*-GAALIE | 1.95 | 3.24 | 0.45 | 10.51 | 2.62 | 2.88 |

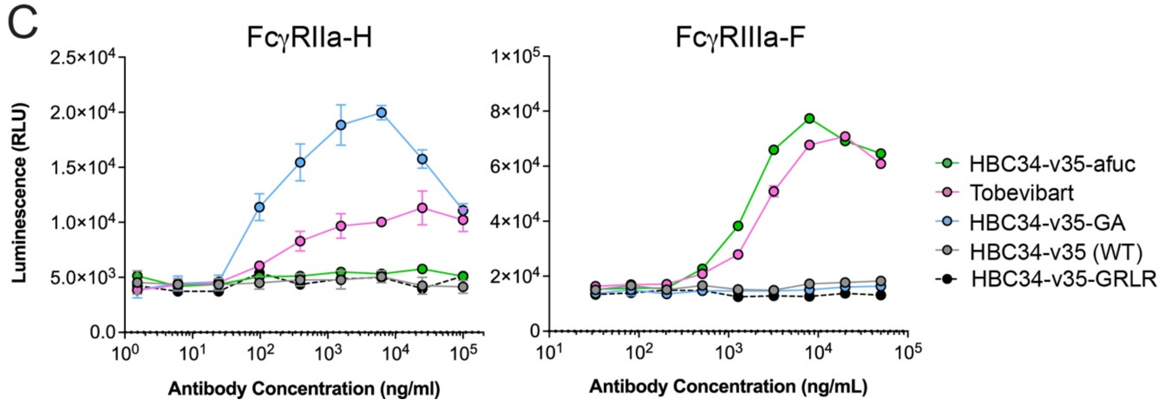

**Supplementary Figure S3: HBC34* Fab and Fc variants bind and activate FcγRs.**
**(A)** Binding of HBC34* Fc variants (single mAbs in solution, all with the LS-Fc mutation) to the indicated immobilized human FcγRs, measured via SPR while stepwise increasing the mAb concentrations.
**(B)** Fold change of $K_D$ values of FcγRs binding of HBC34* Fc variants relative to WT Fc from SPR measurements in (A). Larger values indicate improved binding.
**(C)** HBC34-v35 Fc variants (including tobevibart, which carries the LS-GAALIE Fc) inducing signaling of FcγRIIa H131 (left) or FcγRIIIa F158 (right) using 250 or 5,000 IU/mL recombinant HBsAg (Prospec). Representative results of one out of 2 independent experiments per FcγR.

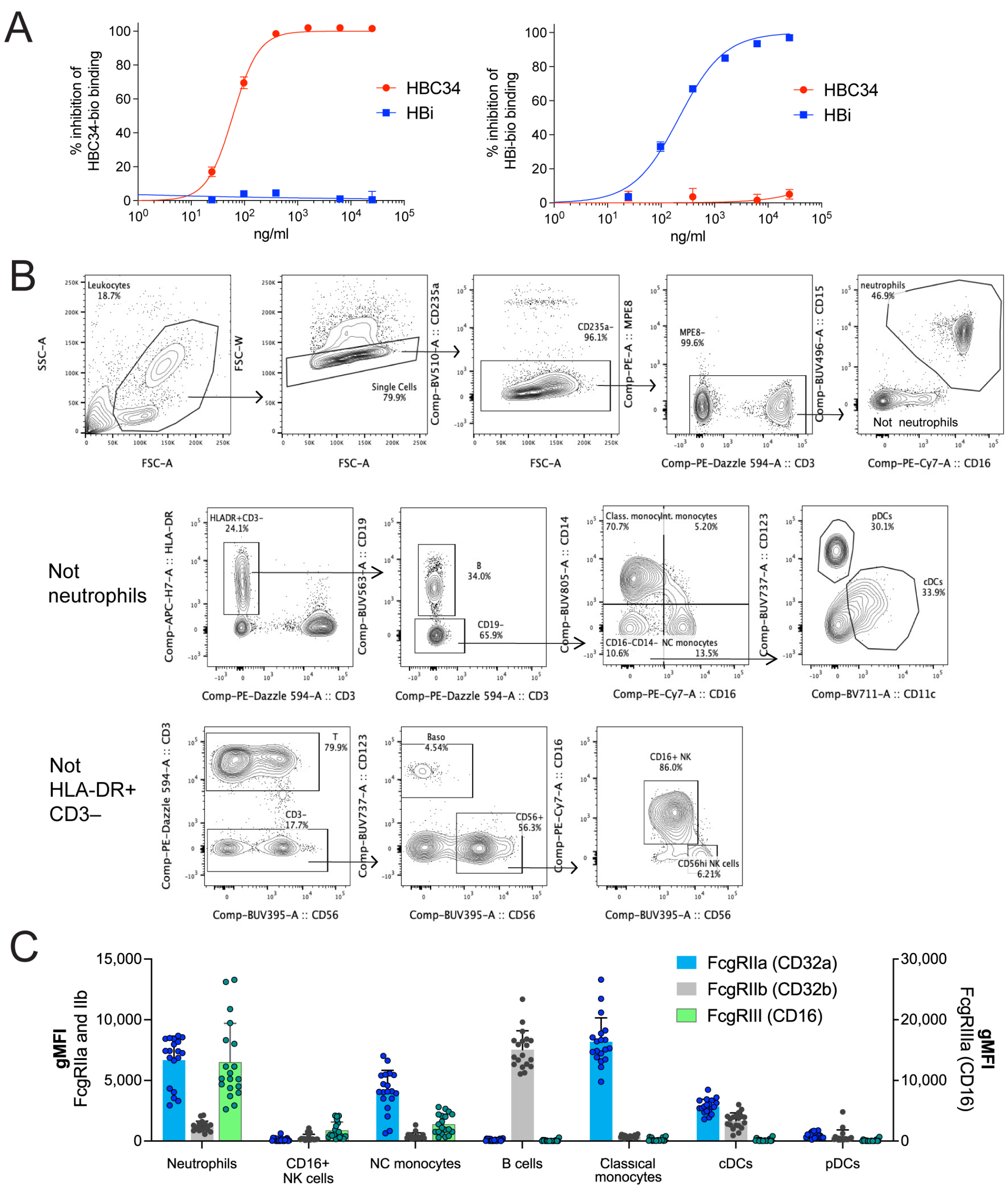

**Supplementary Figure S4: HBi does not compete with HBC34 binding, and gating of immune cell populations in human whole blood following addition of HBC34* Fc variants *in vitro*.**

**(A)** Competition ELISA showing that HBi does not compete with HBC34-bio binding but that HBC34 inhibits its own binding (of HBC34-bio, left). Vice versa, HBC34 does not compete with HBi binding but HBi inhibits its own binding (of HBi-bio, right).

**(B)** Gating of cell populations in whole blood of patients with CHB, based on flow cytometry markers.

**(C)** Bar graph showing FcγRIIa, FcγRIIb, and FcγRIIIa/b expression (means ±SD of gMFI and individual data points) across immune cell populations. Analysis of n=19 of samples from patients with CHB in main Figure 3B and C.

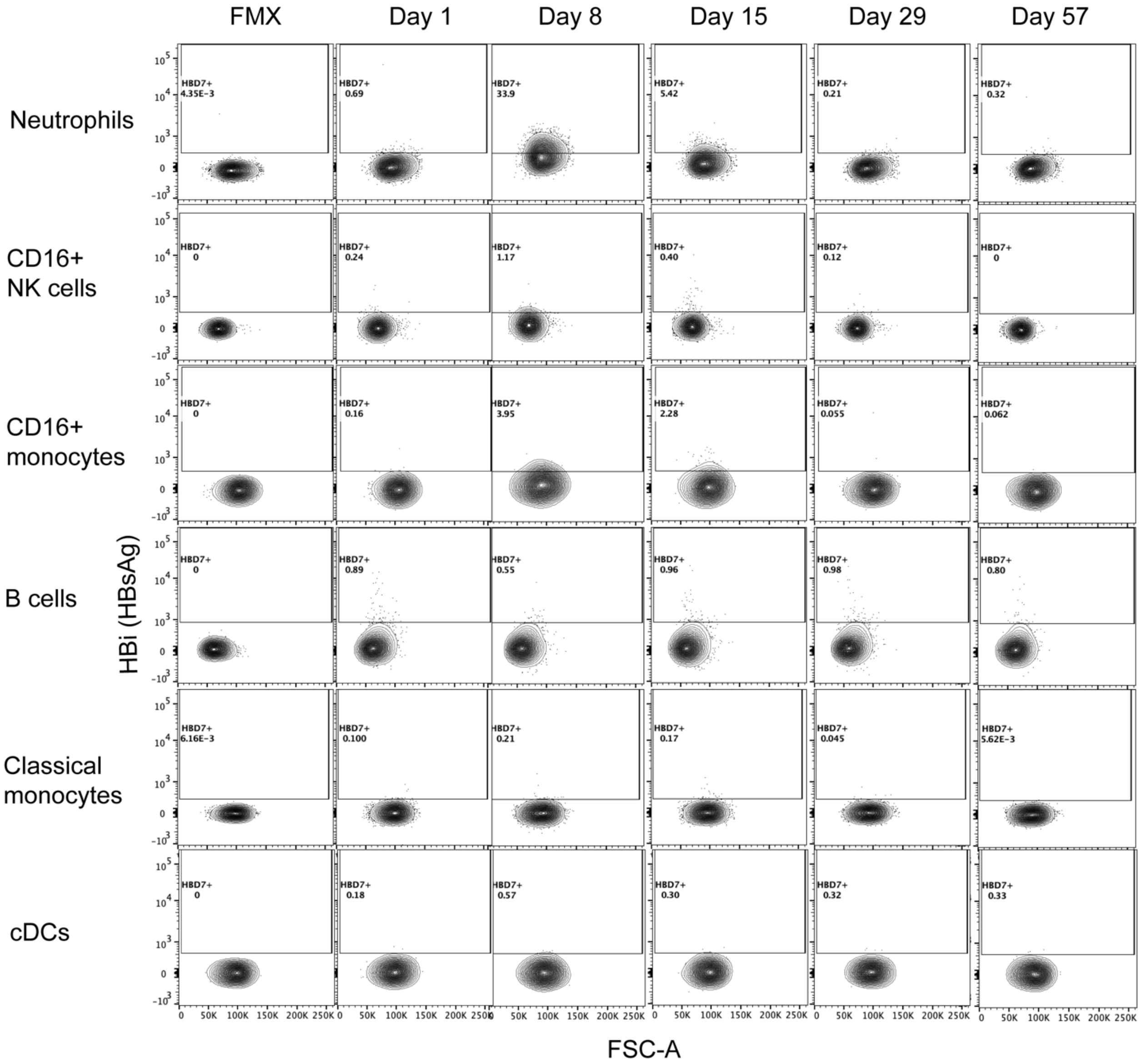

**Supplementary Figure S5: Gating of HBi+ (HBsAg+) cells in samples from patients with CHB dosed with 300 mg of tobevibart in a phase 1 clinical trial.**
Example flow cytometry plots specifying HBi+ (HBsAg+) cells for neutrophils, NC monocytes, CD16+ NK cells, classical monocytes, B cells and cDCs on days 1, 8, 15, 29, and 57 post-dosing of tobevibart. HBi gates were set using the sample prior to dosing tobevibart (day 1) as negative control.

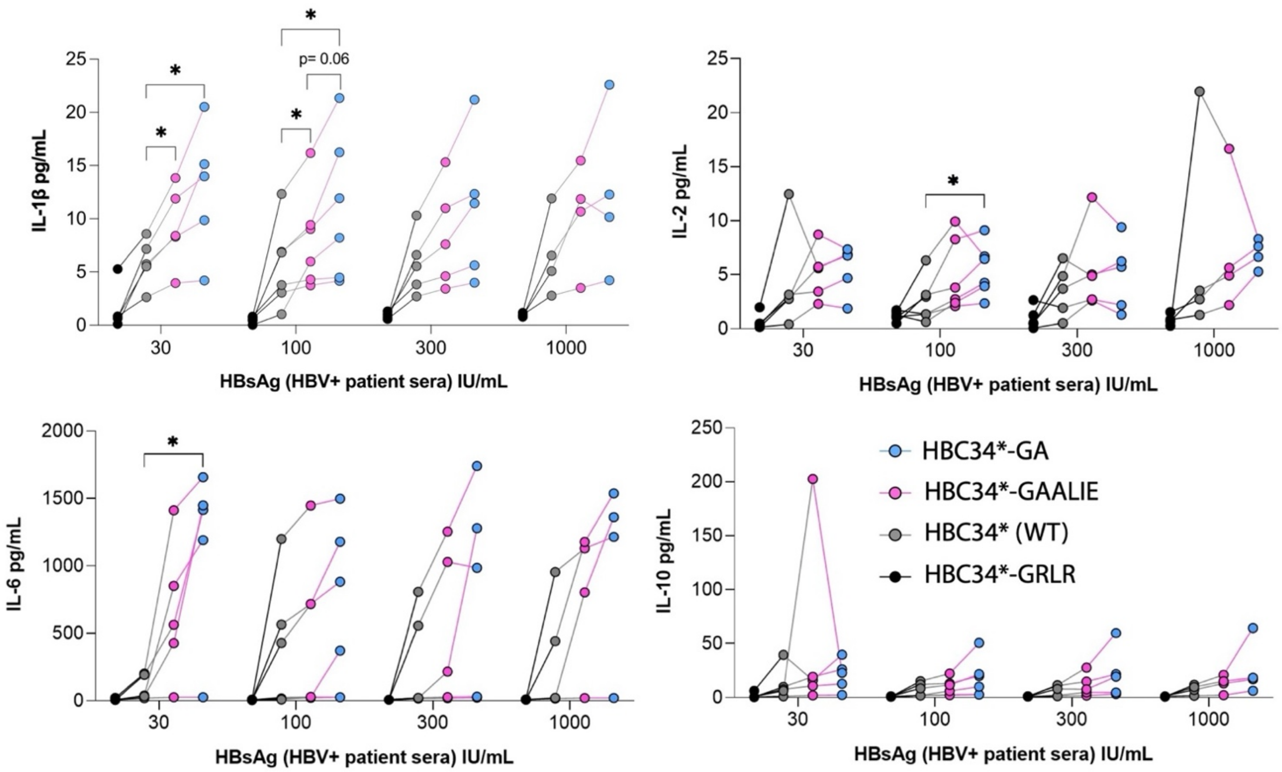

**Supplementary Figure S6: Cytokine secretion of activated human moDCs.**
Summary graphs showing the amount (pg/mL) of IL-1b, IL-2, IL-6, and IL-10 secreted by moDCs stimulated with HBsAg from HBV+ patient sera at 30, 100, 300, 1000 IU/mL in ICs with 50 µg/mL of HBC34*-GAALIE or Fc variants. Results were combined from 3 moDCs donors and 5 HBV+ patient sera. A two-way ANOVA with Geisser-Greenhouse correction and Dunnett's multiple comparison test was used.

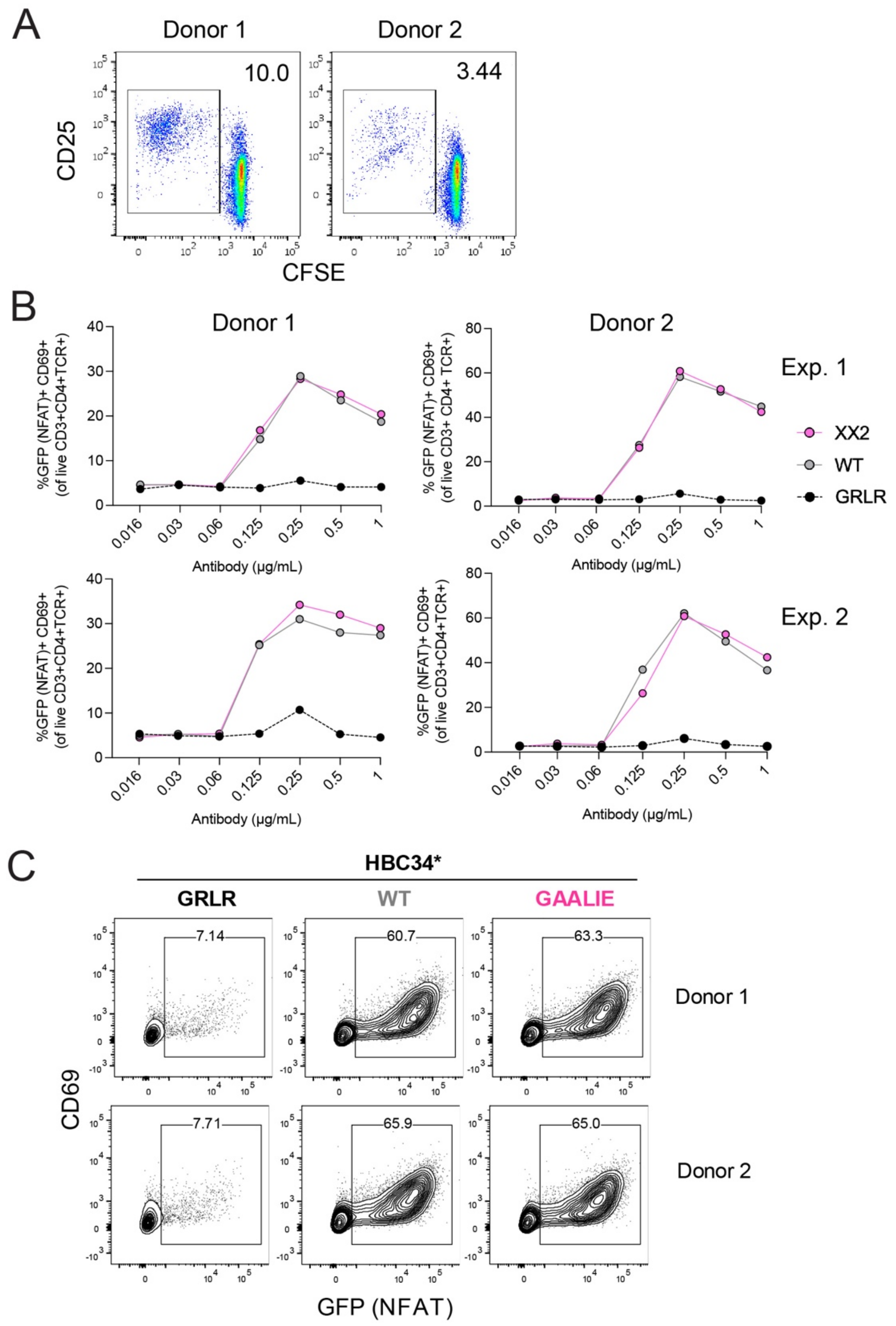

**Supplementary Figure S7: Activation of human CD4+ memory T cells or (TCR)-transgenic Jurkat reporter T cells specific for HBsAg in co-culture with IC-stimulated moDCs.**
**(A)** Flow cytometry plots showing CD25 expression and CFSE dilution of primary human CD4+ memory T cells from HBV vaccinees in co-culture with autologous moDCs of two monocyte donors stimulated only with 100 IU/mL HBsAg as control.
**(B)** Representative graphs of two monocyte donors in 6 independent experiments showing % GFP+ CD69+ activated TCR-transgenic Jurkat reporter cells when in co-culture with HLA-matched moDCs stimulated with HBsAg generated from PLC cells at 1000 IU/mL in complex with different concentrations of HBC34*-GAALIE or Fc variants (GRLR or WT).
**(C)** Representative flow cytometry plots showing the percentage of CD69+ GFP+ reporter T cells when in co-culture with moDCs of two donors stimulated with HBsAg generated from PLC cells at 1000 IU/mL as in **(A)** in complex with 0.25 µg/mL of HBC34*-GAALIE or Fc variants (GRLR or WT).

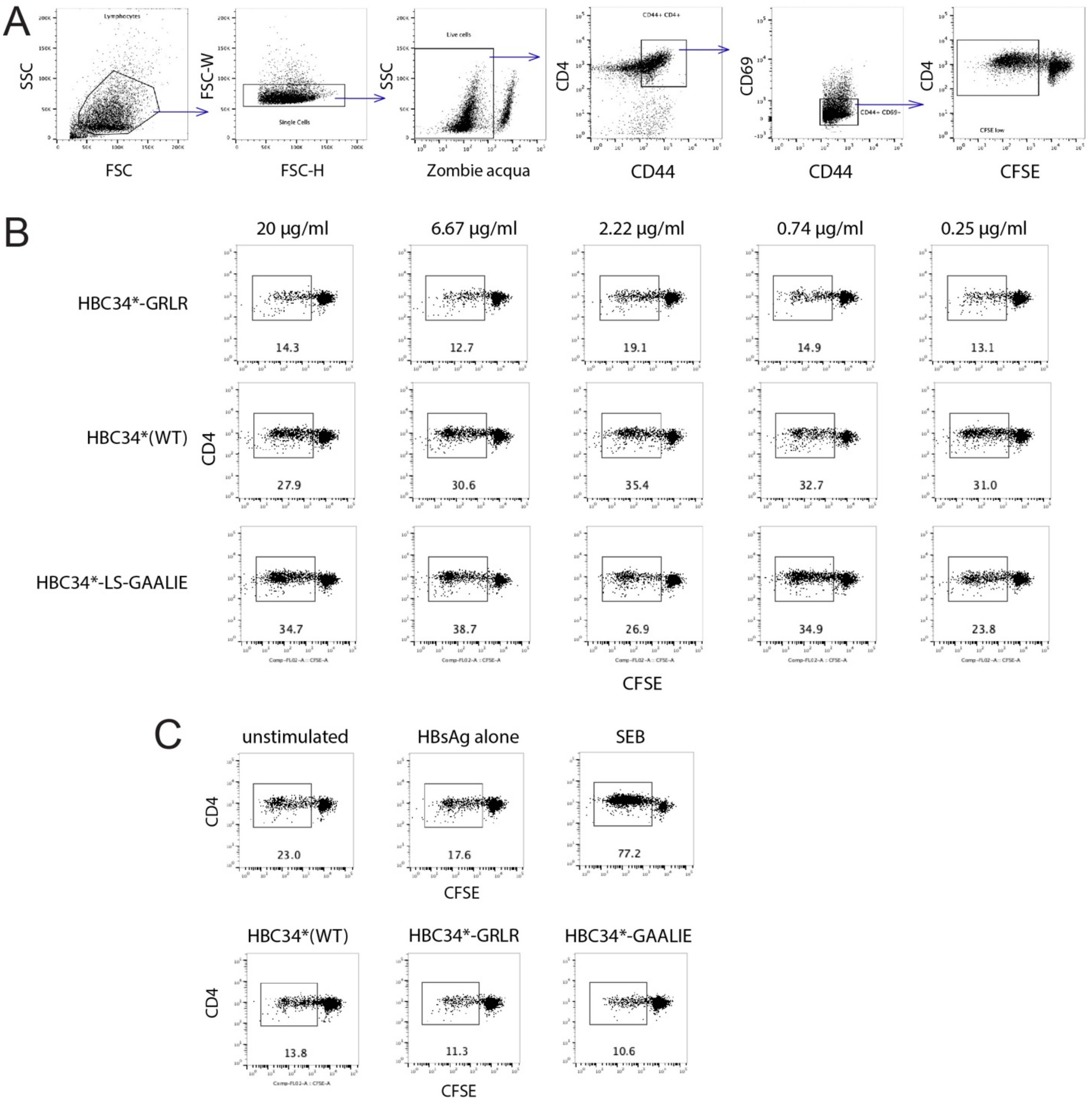

**Supplementary Figure S8: Gating strategy, HBC34*-GAALIE ICs more efficiently activate CD4+ memory T cells of HBV-vaccinated mice transgenic for human FcγRs and.**

**(A**) Gating used to determine proliferation of CD4+ CD44+ CD69- CFSE low memory T cells from HBV-vaccinated mice in flow cytometry data.

**(B)** Representative flow cytometry plots showing % proliferating CD4+ CD44+ CFSE low memory T cells re-stimulated *in vitro* with ICs containing 1,000 IU/mL HBsAg and titrated HBC34* Fc variants (GRLR, WT or GAALIE)

**(C)** Representative flow cytometry plots of control cultures containing medium only (unstimulated), HBsAg alone (1,000 IU/mL), SEB (1 µg/mL), or mAb HBC34* Fc variants alone (20 µg/mL).
